# Aortic Geometric Atlas: Centile-Based Reference Charts and Pathological Signatures Across the Adult Lifespan

**DOI:** 10.64898/2026.06.25.26356450

**Authors:** Cameron A. Beeche, Hamed Tavolinejad, Zhirui Li, Zirui Fan, Bingxin Zhao, Heng Wei, Leonard Steger, Rakesh Sharma, Luke Ni, Saif Zaman, Jeffrey Duda, James Gee, Anurag Verma, Hersh Sagreiya, Scott M. Damrauer, Michael G. Levin, Daniel Rader, Ari Goldberg, Julio A. Chirinos, Walter R. Witschey, Penn Medicine Biobank

**Author notes:** Corresponding author: Cameron A. Beeche, PhD Student South Tower, Rm. 11-155. Perelman Center for Advanced Medicine. 3400 Civic Center Blvd. Philadelphia, PA. 19104. A full list of contributions from Penn Medicine BioBank team is provided in supplement.

## Abstract

The aorta is a site of major cardiovascular burden, yet its clinical assessment on computed tomography (CT) remains limited to manual diameter measurements. Here we present the Aortic Geometric Atlas, a comprehensive characterization of thoracic aortic geometry, realized through the Aortic Geometry Toolkit (AGT), an automated pipeline we developed to extract 38 aortic geometric phenotypes (AGPs) across anatomically delineated subsegments. Applying AGT to 62,366 participants representing 140,319 CT studies, we constructed sex-specific, continuous, centile-based reference ranges spanning nine decades of the adult lifespan from 35,648 participants without aortic disease. We then performed a phenome-wide time-to-event analysis for incident disease, identifying 861 prognostic associations across 155 phecodes, with non-caliber geometry contributing predictive value beyond diameter, and derived disease-specific AGP signatures for cardiovascular risk stratification. Together, the Aortic Geometric Atlas provides a population-scale reference for individualized aortic assessment, positioning AGPs as early subclinical markers of incident cardiovascular disease.

## INTRODUCTION

The thoracic aorta is a major site of cardiovascular pathology, with disease spanning a broad clinical spectrum from highly prevalent atherosclerosis to potentially lethal aneurysm and dissection that together account for over 150,000 deaths globally each year^1–3^. In clinical practice, however, the aorta is assessed almost entirely by a small number of manual diameter measurements, which are observer-dependent and systematically overestimated^4–7^. Cardiac structure, by contrast, is comprehensively characterized with well-established reference values^8–10^. Recently, aortic geometric phenotypes (AGPs^11,12^), including diameter, length, curvature, tortuosity, cross-sectional area and volume, have each demonstrated independent prognostic value for aortic and cardiac events^13–20^. Computed tomography (CT) is among the most widely performed diagnostic imaging modalities, with over 300 million scans acquired globally each year^21–24^, yet this rich source of three-dimensional aortic data remains largely unleveraged. Moreover, population-level references have been established for aortic diameter across several cohorts^25–28^, but these efforts have relied on manual measurements subject to the same biases, and have not been translated into continuous, centile-based references across the adult lifespan. Although deep learning has enabled automated aortic segmentation^29^ and AGP quantification, no unified atlas of aortic geometry currently exists to enable individualized contextualization against substantial reference aging trajectories. To address this gap, we curated the largest retrospective, longitudinal, multi-institutional dataset for thoracic aortic imaging to date, comprising over 70 distinct scanners, 63,920 participants, 147,167 studies, and 393,948 CT volumes from the Penn Medicine Biobank (PMBB) and CT-RATE datasets^30,31^. We propose the Aortic Geometric Atlas, realized through the Aortic Geometry Toolkit (AGT), an open-source, publicly available pipeline we developed that automatically extracts comprehensive regional aortic geometry across anatomically delineated subsegments of the thoracic aorta. From this dataset, we identify reference participants without documented aortic disease and construct sex-specific aortic aging charts spanning nine decades of the adult lifespan, exceeding all prior aortic reference studies in both sample size and age range. We score participants with documented aortic pathology, including atherosclerosis, aneurysm, dissection, and coarctation, against this reference to characterize the centile profiles of each condition across all AGPs. We then perform a phenome-wide time-to-event analysis for incident disease, demonstrating that non-caliber phenotypes including length, tortuosity, and branch vessel architecture carry independent predictive value beyond diameter. Finally, we derive disease-specific AGP signatures for cardiovascular risk stratification. Together, the Aortic Geometric Atlas establishes a population-scale foundation for individualized aortic assessment and a pathway toward routine use of AGPs as subclinical markers of cardiovascular risk.

## RESULTS

We assembled a retrospective multi-institutional dataset from the PMBB and CT-RATE, comprising 63,920 participants, 147,167 imaging studies, and 393,948 CT volumes (**Table S1**). The PMBB cohort included 42,621 adults who underwent thoracic CT imaging, comprising 121,482 studies and 343,777 CT volumes, with a median age of 60.9 years (IQR=49.6-69.4), balanced sex distribution (49.6% male, 50.4% female), and representation across White (73.1%), Black (19.6%), Asian (2.5%), and other (4.8%) racial groups. The CT-RATE dataset included 21,299 participants with 25,685 studies and 50,171 CT volumes (median age=45.0 years, IQR=34.0-61.0, 57.5% male). In PMBB, 89,733 studies (73.9%) had an associated radiology report; all CT-RATE studies had associated reports. Full demographic characteristics are provided in **Table S1** and scanner acquisition parameters, including manufacturer, pixel spacing, and slice thickness, are provided in **Table S2**.

### Aortic geometry toolkit

To enable comprehensive geometric characterization of the aorta from CT, we developed the AGT, a computational pipeline that transforms a three-dimensional aortic segmentation into a structured set of regional geometric phenotypes (**Fig. 1**). The pipeline begins with segmentation of the thoracic aorta using TotalSegmentator^29^, followed by inferior cropping at the T12 vertebral level to isolate the thoracic segment. From the aorta segmentation, a smoothed triangular surface mesh is generated via marching cubes, and an ordered centerline is extracted through 3D skeletonization, graph-based longest-path traversal, and cubic spline fitting. Anatomical branch vessel landmarks, specifically the brachiocephalic trunk and left subclavian artery, are automatically detected from the multi-label segmentation and used to partition the centerline into the ascending aorta, aortic arch, and descending aorta. Extracted AGPs are organized into three hierarchical orders reflecting the level of abstraction from the underlying centerline. First-order phenotypes are direct geometric measurements, including diameter, cross-sectional area, centerline length, volume, surface area, arch height, arch width, and branch vessel takeoff angles at the brachiocephalic trunk, left common carotid, and left subclavian arteries. Second-order phenotypes are derived through differentiation or ratios of first-order quantities and include curvature, tortuosity, and taper ratio. Third-order phenotypes capture local cross-sectional shape or higher-order centerline geometry and include eccentricity and torsion. In total, the AGT extracts 38 distinct AGPs per CT volume. Full details are provided in **Methods**.

**Figure 1.**
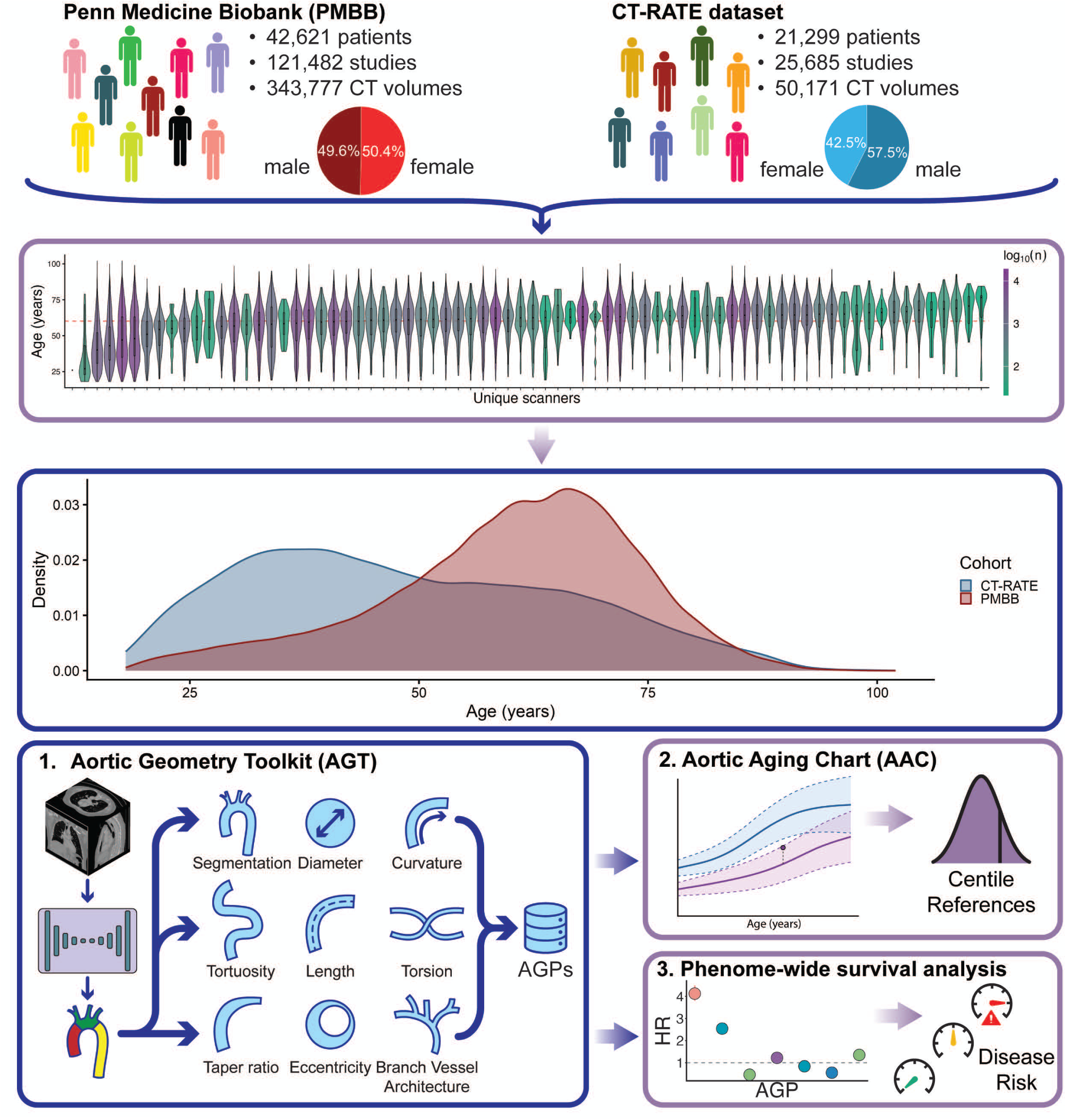
Study overview and the Aortic Geometry Toolkit. A multi-institutional dataset was assembled from the Penn Medicine Biobank (PMBB; 42,621 participants, 121,482 studies, 343,777 CT volumes; 50.4% female) and the CT-RATE dataset (21,299 participants, 25,685 studies, 50,171 CT volumes; 42.5% female), spanning over 70 unique scanner models with complementary age distributions across the adult lifespan. The Aortic Geometry Toolkit (AGT) extracts 38 aortic geometric phenotypes (AGPs) across nine categories (diameter, cross-sectional area, length, curvature, tortuosity, torsion, eccentricity, taper ratio, and branch vessel architecture) from anatomically delineated subsegments defined by brachiocephalic trunk and left subclavian artery landmarks. AGPs from a reference cohort free of documented aortic disease are used to construct sex-specific Aortic Aging Charts (AAC) via GAMLSS modeling, producing continuous centile-based reference curves across the adult lifespan. The prognostic utility of AGPs is evaluated through a phenome-wide time-to-event analysis for incident disease and disease-specific AGP signature models, demonstrating independent predictive value beyond conventional diameter measurements.

We applied the AGT across both cohorts, successfully extracting AGPs from 140,319 of 147,167 studies (95.3%) spanning 62,366 of 63,920 participants (97.6%) (**Table S1**). The remaining failures occurred primarily due to incomplete fields of view that prevented the segmentation from encompassing the complete thoracic aorta. Representative examples of AGT phenotype extraction across various contrast-protocols, fields of view, and pathological conditions are provided in **Figures S1-S8**.

### Aortic aging charts

To construct reference ranges for aortic geometry spanning the adult lifespan (ages 18-99), we identified 59,588 studies from 35,648 participants without documented aortic diseases (20,703 from PMBB and 14,945 from CT-RATE; **Table S3**). Adapting WHO recommendations for growth charts, we modeled each AGP as a nonlinear function of age using generalized additive models for location, scale, and shape (GAMLSS)^32,33^, with sex included as an interaction term on the location and scale parameters to capture sex-specific trajectories, while cohort was incorporated as a random effect to account for cohort-level variation. Following previous work^34,35^, we performed automated model selection across 9 candidate specifications, comprising three fractional polynomial orders for the location and scale parameters crossed with three distribution families (**Methods**). To confirm stability of the fitted reference curves against sampling variation, we performed a stratified 50-50 discovery-replication split of the reference cohort and independently refit each chart on the two halves, observing agreement in median centile curves (mean female R^2^=0.983, male R^2^=0.990; **Supplemental Materials**, **Table S4**, and **Figs. S9-S10**). As a sensitivity analysis, we replaced the cohort-level random effect with a scanner x cohort random effect (74 levels; **Methods**) and compared the resulting centile curves, observing high agreement across all phenotypes (mean female R^2^=0.982, male R^2^=0.972; 71 of 76 pairs R^2^ > 0.95; **Table S5, Figs. S11-S12**), suggesting that the fitted reference curves are robust to scanner-level variation.

### Global aortic aging patterns

Aortic aging charts revealed distinct, nonlinear trajectories of geometric remodeling across the adult lifespan (**Fig. 2**). Growth curves exhibited characteristic patterns, with sex-specific effects that were predominantly linear. Total thoracic aortic volume increased approximately 2.5-fold over the adult lifespan in both sexes, from a median of 113 mL in males and 84 mL in females at age 18 to 290 mL in males and 212 mL in females by ages 97 and 93, respectively (**Fig. 2a**). Aortic diameter increased steadily throughout most of the adult lifespan, reaching a maximum at approximately age 86 in both sexes (male 95% CI=84.3, 87.7; female 95% CI=83.8, 87.3), after which it exhibited modest regression (**Fig. 2b**). Overall diameter grew from 1.83 cm to 2.70 cm in males and 1.64 cm to 2.42 cm in females between ages 18 and 86. Females showed consistently smaller diameters but comparable growth rates. In contrast to diameter, thoracic aortic length grew continuously throughout the entire adult lifespan (**Fig. 2c**). Centerline length in males increased from 28.9 cm to 36.9 cm between ages 18 and 99, representing a 28% elongation; females showed a concordant trajectory from 26.3 cm to 33.7 cm. Notably, the rate of elongation itself accelerated with age, increasing from 0.087 cm/year at age 18 to 0.111 cm/year at age 99 in males (0.081 to 0.104 cm/year in females), indicating that the aorta elongates progressively faster as it ages.

**Figure 2.**
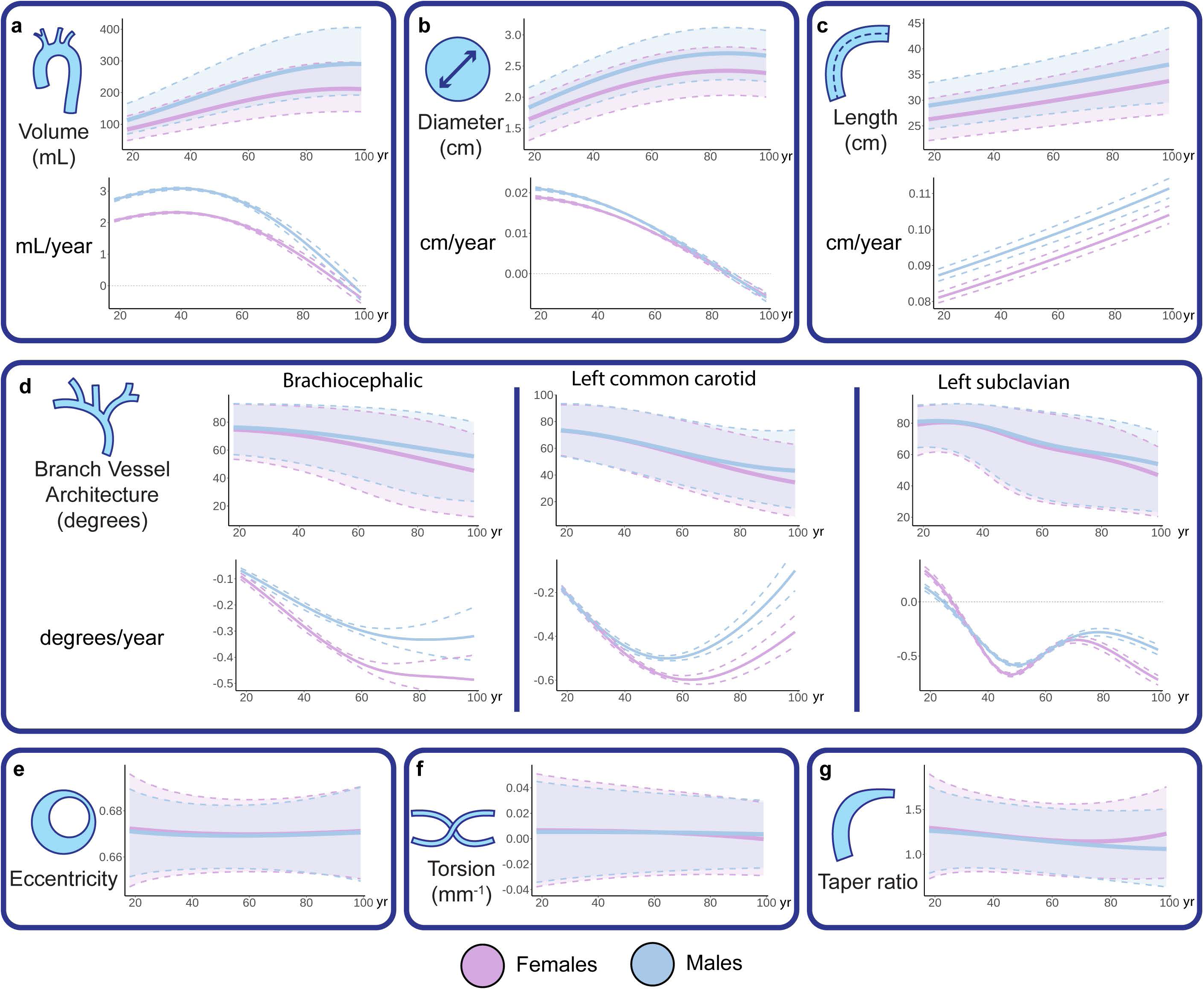
Global aortic aging patterns across the adult lifespan. Sex-specific aortic aging charts (blue, male; pink, female; solid lines, median; shaded regions, 2.5th–97.5th centiles) with corresponding first derivatives (rates of change) for global aortic geometric phenotypes. (**a**) Total thoracic aortic volume increased approximately 2.5-fold over the adult lifespan in both sexes, with the rate of volumetric growth declining monotonically. (**b**) Overall aortic diameter increased steadily throughout most of adulthood, reaching a maximum at approximately age 86 before exhibiting modest regression; the growth rate declined continuously, crossing zero in the ninth decade. (**c**) Overall centerline length increased continuously throughout the entire adult lifespan with no plateau observed, and the rate of elongation itself accelerated with age. (d) Branch vessel takeoff angles for the brachiocephalic trunk, left common carotid, and left subclavian arteries declined progressively with age, with females exhibiting a more rapid decline; the left subclavian showed a delayed onset of remodeling relative to the more proximal vessels. Rates of angular decline are shown below each branch vessel. (e) Aortic eccentricity, (**f**) torsion, and (**g**) taper ratio remained essentially static across the adult lifespan, with no meaningful age-dependent change in either sex.

Branch vessel geometry also showed age-dependent remodeling (**Fig. 2d**). The brachiocephalic trunk takeoff angle began at nearly identical values between the sexes at age 18 (76.3° in males and 74.7° in females), then diverged with age, declining to 55.5° in males and 45.2° in females by age 99. The left common carotid artery showed a concordant pattern. The left subclavian artery exhibited similar remodeling but with a delayed onset, declining from 81.4° in males and 80.6° in females to 53.9° and 46.8°, respectively, by age 99. This delayed remodeling may reflect its more distal position along the arch relative to the aortic root. This progressive reduction in branch vessel takeoff angles is consistent with aortic elongation and unfolding, as the branch vessel origins shift relative to the arch apex toward the ascending aorta with age^36^. Aortic eccentricity, torsion, and taper ratio remained essentially static across the adult lifespan, with no meaningful change in either sex (**Figs. 2e-g**). Specifically, aortic eccentricity remained stable at approximately 0.67 (aortic eccentricity index=0.33). Longitudinal assessment of paired studies demonstrated a reproducibility gradient tracking phenotype complexity, with first-order phenotypes showing the highest stability and third-order phenotypes the lowest. Complete results are provided in **Supplemental Results**, **Figs. S13-S24** and **Table S7**.

### Regional heterogeneity of aortic remodeling

Aging charts constructed for each thoracic aortic subsegment revealed substantial heterogeneity in remodeling patterns (**Fig. 3**). The ascending aorta dilated continuously throughout adulthood, with median diameter increasing from 2.10 cm in males and 1.91 cm in females at age 18 to 2.96 cm and 2.70 cm by age 83 (**Fig. 3a**). The arch and descending aorta exhibited similar patterns but with smaller absolute diameters (arch: 2.76 cm males, 2.54 cm females; descending: 2.61 cm males, 2.31 cm females). Aortic length increased continuously throughout adulthood across all subsegments (**Fig. 3b**). Tortuosity exhibited regional heterogeneity across subsegments (**Fig. 3c**), with ascending tortuosity remaining essentially constant, arch tortuosity decreasing nearly three-fold by age 78, and descending tortuosity increasing over three-fold with the rate of increase peaking at age 64 in males and 67 in females. Similarly, curvature exhibited heterogeneous patterns across subsegments (**Fig. 3d**), with ascending and arch curvature declining monotonically while descending aortic curvature increased progressively. Complete aging chart results for all AGPs are provided in **Table S6**.

**Figure 3.**
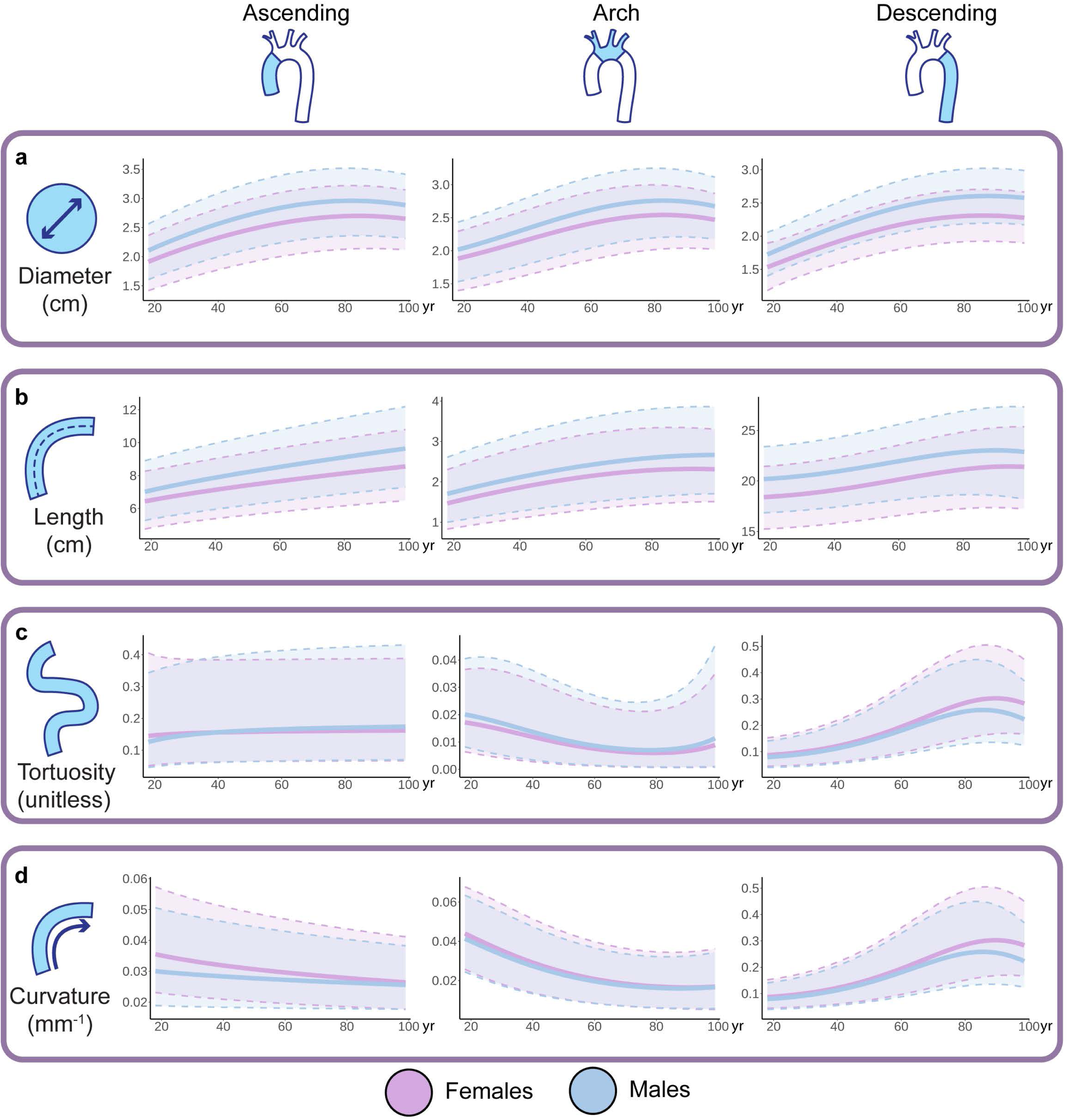
Regional heterogeneity of aortic remodeling across the adult lifespan. Sex-specific aortic aging charts for four geometric phenotypes across the ascending aorta, aortic arch, and descending aorta (blue, male; pink, female; solid lines, median; dashed lines, 2.5th and 97.5th centiles). **(a)** Diameter increased throughout most of adulthood across all subsegments, reaching a maximum at approximately age 83-88 before exhibiting modest regression, with more proximal segments plateauing earlier than distal segments. **(b)** Centerline length increased continuously throughout the entire adult lifespan across all subsegments, with no plateau observed, and the rate of elongation accelerated with age. **(c)** Tortuosity exhibited the most striking regional heterogeneity: ascending aorta tortuosity remained essentially static, arch tortuosity decreased nearly three-fold, while descending aorta tortuosity increased over three-fold across the adult lifespan. **(d)** Curvature showed monotonic decline in the ascending aorta and arch, consistent with progressive arch flattening, while the descending aorta exhibited a U-shaped trajectory, declining through early adulthood before progressively increasing. Together, these charts reveal that the thoracic aorta undergoes a gradual transformation from a compact arch to an elongated, tortuous configuration, with the timing and magnitude of remodeling varying substantially across subsegments.

### Centile profiles of aortic pathology

We scored participants with documented aortic pathology against the AAC reference to quantify condition-specific patterns of geometric deviation (**Methods**). Across the four conditions, 117 of the 152 AGP comparisons deviated significantly from the AAC reference (*P* < 3.29×10^−4^). Atherosclerotic aortas (n=19,180) exhibited modestly elevated caliber centiles (overall diameter median centile=0.65; IQR=0.36-0.87), with non-caliber AGPs remaining approximately normative (**Fig. 4a**). Aneurysmal aortas (n=2,263) showed caliber elevations, evidenced by volume (median centile=0.98; IQR=0.89, 1.00) and overall diameter (median centile=0.95; IQR=0.80-0.99) (**Fig. 4b**). Moreover, aneurysm participants exhibited secondary elevations in centerline length (median centile=0.83; IQR=0.56-0.97) and descending tortuosity (median centile=0.77; IQR=0.42-0.96), suggesting that the aneurysmal geometric distortion extends beyond caliber alone. Dissected aortas (n=274) exhibited AGP elevations concentrated in the descending aorta, with diameter (median centile=0.99), tortuosity (median centile=0.97), and curvature (median centile=0.95) all markedly elevated, alongside global increases in volume (median centile=0.99) and eccentricity (median centile=0.93) (**Fig. 4c**), reflecting the severe geometric distortion characteristic of dissection. Coarctation participants (n=44) exhibited a bimodal centile profile, with arch diameter severely reduced (median centile=0.045) alongside elevations in overall curvature (median centile=0.93), descending curvature (median centile=0.91), and arch tortuosity (median centile=0.90) (**Fig. 4d**). Complete centile profiles are presented in **Table S8**. Additionally, we trained logistic regression models on the PMBB to predict prevalent aortic diseases (aneurysm, atherosclerosis, and dissection) indicated in the radiology reports and performed external testing on CT-RATE (**Methods**). In the PMBB cross-validation, AGPs predicted aortic aneurysm with an AUROC of 0.917 (95% CI=0.914, 0.920), whereas atherosclerosis yielded a lower AUROC of 0.714 (95% CI=0.712, 0.716). Finally, dissection was predicted with an AUROC of 0.948 (95% CI=0.943, 0.954). Performance generalized to the external CT-RATE validation, with AUROCs of 0.916 (95% CI=0.907, 0.925) for aneurysm, 0.861 (95% CI=0.856, 0.867) for atherosclerosis, and 0.981 (95% CI=0.959, 1.000) for dissection (**Figs. 4e-g**). Complete results are presented in **Table S9**.

**Figure 4.**
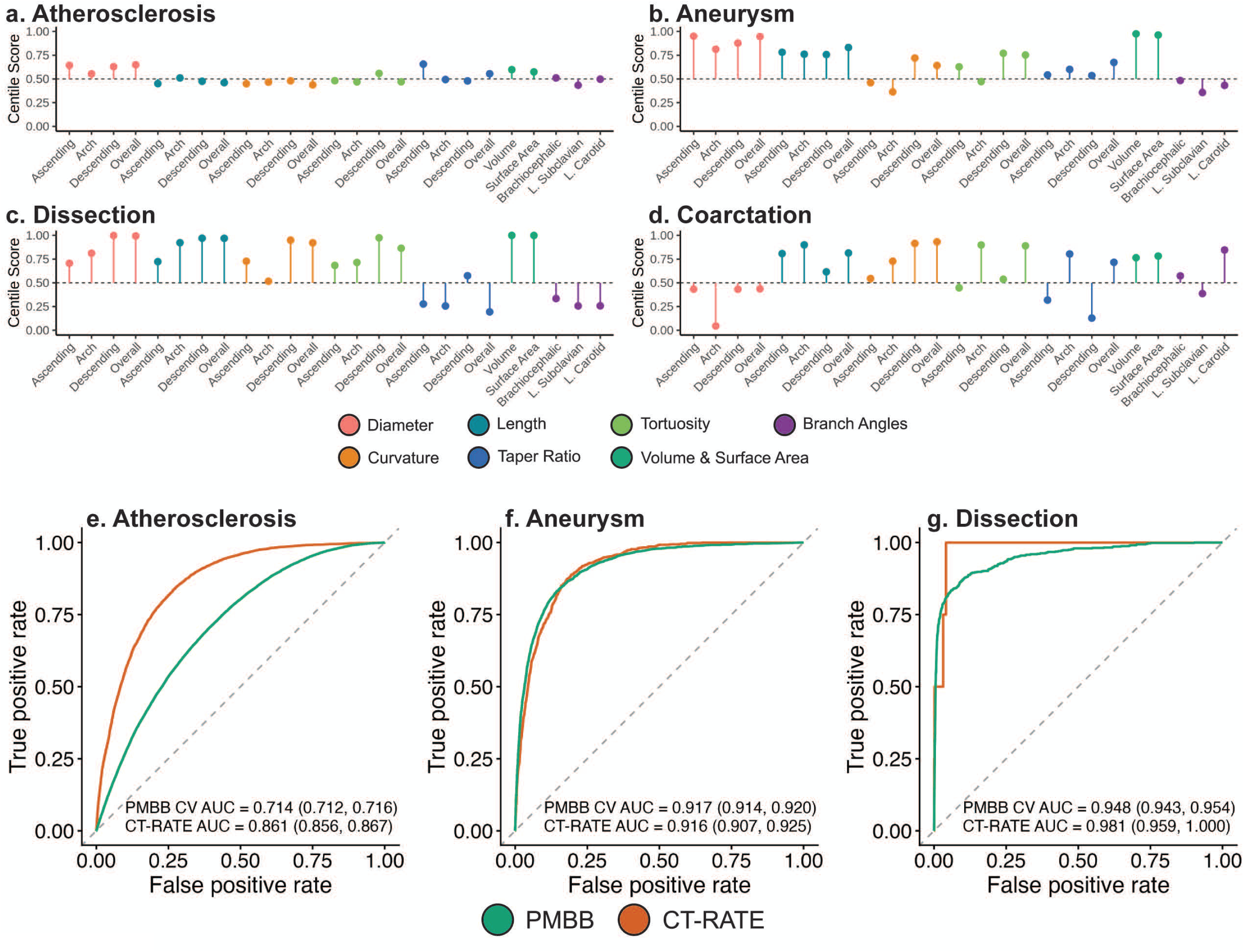
Centile profiles of aortic pathology. Median centile scores across select aortic geometric phenotypes (AGPs) for participants with documented (**a**) atherosclerosis, (**b**) aneurysm, (**c**) dissection, and (**d**) aortic coarctation, scored against aortic aging charts. Each lollipop represents the per-AGP median centile, colored by phenotype category, with the dashed line indicating the median (0.50). Atherosclerosis exhibited modest, predominantly caliber-centered elevations. Aneurysm showed dramatic caliber elevations with secondary increases in length and tortuosity. Dissection displayed the most extreme centile shifts, with near saturation of caliber phenotypes and marked elevations in descending aorta tortuosity and curvature, accompanied by reduced branch vessel angles. Coarctation exhibited a distinctive bimodal profile combining severely reduced arch diameter (median centile 0.045) with markedly elevated curvature and tortuosity, reflecting the geometric distortion required to accommodate the stenotic segment. These condition-specific geometric signatures demonstrate that aortic pathologies produce distinct patterns of deviation across the full AGP space that would be invisible to diameter-only assessment. (**e-g**) External validation of AGP-based disease classification. Logistic regression models were trained in the PMBB using AGPs selected by reverse stepwise elimination, then externally tested in CT-RATE. Receiver operating characteristic curves are shown for (**e**) atherosclerosis, (**f**) aneurysm, and (**g**) dissection, each comparing internal PMBB cross-validation against external CT-RATE testing, with the area under the curve (AUROC).

### Prognostic utility of aortic geometry across the disease phenome

The phenome-wide time-to-event analysis (**Fig. 5a**) identified 861 significant associations (*P* < 1.018×10^−6^) across 155 phecodes and 28 AGPs, of which 318 associations (spanning 106 phecodes) involved non-caliber phenotypes including length, tortuosity, curvature, eccentricity, taper ratio, and branch vessel architecture (**Fig. 5b**). Aortic aneurysm exhibited the strongest associations, with 21 significant AGPs spanning 7 phenotype categories (**Fig. 5c**). Ascending aortic diameter showed the largest effect (HR=4.16; 95% CI=3.81, 4.53), but 11 significant associations involved non-caliber phenotypes including centerline length (ascending HR=2.53 (95% CI=2.35, 2.73); overall HR=2.29 (95% CI=2.15, 2.44)), overall tortuosity (HR=1.71; 95% CI=1.62, 1.81), and overall taper ratio (HR=1.17; 95% CI=1.13, 1.22). Atrial fibrillation was predicted by 11 AGPs (**Fig. 5d**) including three centerline length measurements (overall HR=1.32 (95% CI=1.25, 1.39); ascending HR=1.18 (95% CI=1.11, 1.25); descending HR=1.22 (95% CI=1.19, 1.33)), consistent with prior work linking aortic elongation and arch remodeling to increased left ventricular mass and adverse ventricular-atrial coupling^37^. Incident hypertension was predicted by 11 AGPs (**Fig. 5e**), combining increased descending aortic diameter (HR=1.24; 95% CI=1.18, 1.30), volume (HR=1.28; 95% CI=1.23, 1.33), overall length (HR=1.14; 95% CI=1.09, 1.19), and brachiocephalic angle (HR=1.15; 95% CI=1.11, 1.20) with reduced arch height (HR=0.85; 95% CI=0.82, 0.88) and tortuosity (HR=0.88; 95% CI=0.84, 0.92), consistent with pressure-mediated aortic enlargement and arch unfolding previously described in hypertensive remodeling^37,38^. Both heart failure with preserved ejection fraction (HFpEF) and reduced ejection fraction (HFrEF) were associated with broadly similar aortic geometric profiles, combining increased caliber (descending diameter, volume), elevated branch vessel angles, and reduced arch height and overall tortuosity (**Fig. S25**). HFpEF exhibited stronger associations with arch height reduction (HR=0.72; 95% CI=0.67, 0.76 vs HR=0.86; 95% CI=0.80, 0.93) and branch vessel angle elevation (brachiocephalic HR=1.32; 95% CI=1.24, 1.40 vs HR=1.17; 95% CI=1.09, 1.25), consistent with the contribution of arterial stiffening and ventricular-arterial coupling to diastolic dysfunction^39–42^. Complete time-to-event results are provided in **Table S10**.

**Figure 5.**
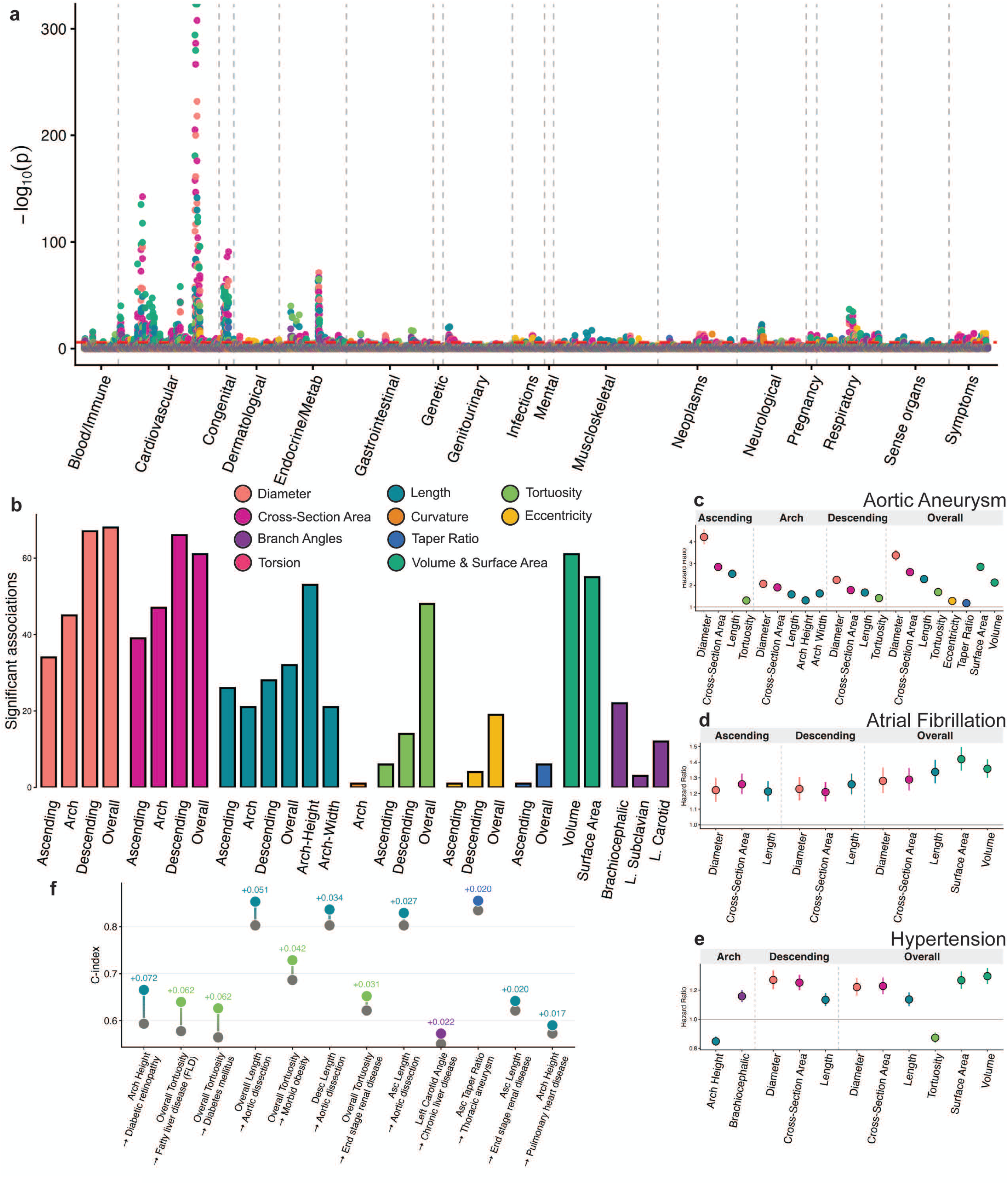
Phenome-wide prognostic associations of aortic geometric phenotypes (AGPs). (**a**) Manhattan plot of phenome-wide time-to-event associations, with each point representing a significant AGP x phecode association plotted by disease group. The y-axis shows −log_10_(P) from Cox proportional hazards models adjusted for age, sex, and age × sex interaction. Points are colored by AGP category. The red line indicates the Bonferroni significance threshold (*P* < 1.018×10^−6^). (**b**) Number of significant phenome-wide associations per AGP, colored by phenotype category, demonstrating that non-caliber phenotypes including length, tortuosity, and branch vessel angles contribute substantial prognostic signal beyond diameter and cross-sectional area. (**c**) Forest plot of hazard ratios with 95% confidence intervals (CI) for all AGPs significantly associated with incident aortic aneurysm, organized by aortic subsegment. Associations span 7 phenotype categories, with 11 involving non-caliber phenotypes. (**d**) Forest plot for incident atrial fibrillation, demonstrating associations across diameter, cross-sectional area, length, and volume phenotypes, with centerline length contributing three independent associations. (**e**) Forest plot for incident hypertension, showing a multi-phenotype signature combining increased caliber, length and branch vessel angles with reduced arch height and tortuosity, characteristic of pressure-mediated aortic enlargement and arch unfolding. (**f**) Independent prognostic value of non-caliber AGPs beyond diameter. Each column shows the baseline C-index (age + sex + age-sex interaction + four regional aortic diameters) and full-model C-index (baseline + non-diameter AGP) for a representative AGP x phecode pair.

To identify AGPs with prognostic value independent of aortic caliber-based measurements, we adjusted for four regional diameter measurements and assessed the incremental change in concordance index (ΔC-index) among AGPs that exhibited significant associations with disease (*P* < 1.382×10^−6^) (**Fig. 5f**). Non-caliber AGPs provided independent discriminative value across 111 phecodes, with notable gains observed across metabolic and vascular diseases including arch height in diabetic retinopathy (ΔC-index+0.072), overall tortuosity in fatty liver disease (ΔC-index+0.062) and diabetes mellitus (ΔC-index+0.062), and overall length in aortic dissection (ΔC-index+0.051). Complete results are presented in **Table S11**.

### AGP signatures

Dimensionality reduction of the AGP space revealed that the two principal axes of geometric variation aligned strongly with age (**Fig. 6a**), consistent with the aging trajectories observed in the AAC. Overlaying disease labels onto the embedding revealed focal clustering of patients with aortic aneurysm in a distinct geometric neighborhood (**Fig. 6b**), suggesting that aneurysmal disease occupies a well-defined region of the aortic geometric phenospace. Disease-specific AGP signatures were derived from multivariate Cox proportional hazards models and benchmarked against baseline and caliber-based alternatives (**Methods**). Of 144 cardiovascular diseases evaluated, 143 yielded an AGP signature. The remaining disease had no AGP associations, leaving it with baseline and caliber models only. Across these 143 cardiovascular diseases, AGP signatures exhibited a mean validation C-index of 0.65 (range=0.51, 0.88) compared with 0.64 (range=0.51, 0.88) for caliber and 0.62 (range=0.49, 0.85) for baseline. A clear performance gradient emerged, where AGP signatures showed marked gains on vascular diseases that tapered across other cardiovascular conditions. For aortic dissection, the AGP signature reached nominal significance (*P* = 0.01), with a validation C-index of 0.86 (95% CI=0.81, 0.91) compared with 0.81 (95% CI=0.76, 0.86) for caliber. Following FDR correction of the 5-year time-dependent AUROC comparison, the AGP signature model for aortic aneurysm (**Fig. 6c**) achieved a validation C-index of 0.86 (95% CI=0.84, 0.87), showing significant performance gains compared with the caliber (C-index=0.82; 95% CI=0.80, 0.84) and baseline (C-index=0.69; 95% CI=0.67, 0.70) models. For aortic valve disorders, the AGP signature reached a validation C-index of 0.74 (95% CI=0.73, 0.76), compared with caliber (C-index=0.72; 95% CI=0.70, 0.74) and baseline (C-index=0.68; 95% CI=0.67, 0.70). Significant AGP signatures also emerged for traditional cardiovascular phenotypes, with more modest gains in discrimination. The AGP signature reached a validation C-index of 0.71 (95% CI=0.70, 0.72) for atrial fibrillation and flutter, 0.66 (95% CI=0.65, 0.67) for hypertension, as well as 0.66 (95% CI=0.65, 0.67) for heart failure. Risk stratification by the AGP signature yielded significant separation (*P* < 3.50×10^−4^) in 113 of 143 conditions, with a mean high-risk enrichment of 2.55-fold (range=1.05x, 22.97x) (**Fig. 6d**). Full comparisons against the caliber and baseline models are provided in **Table S12**.

**Figure 6.**
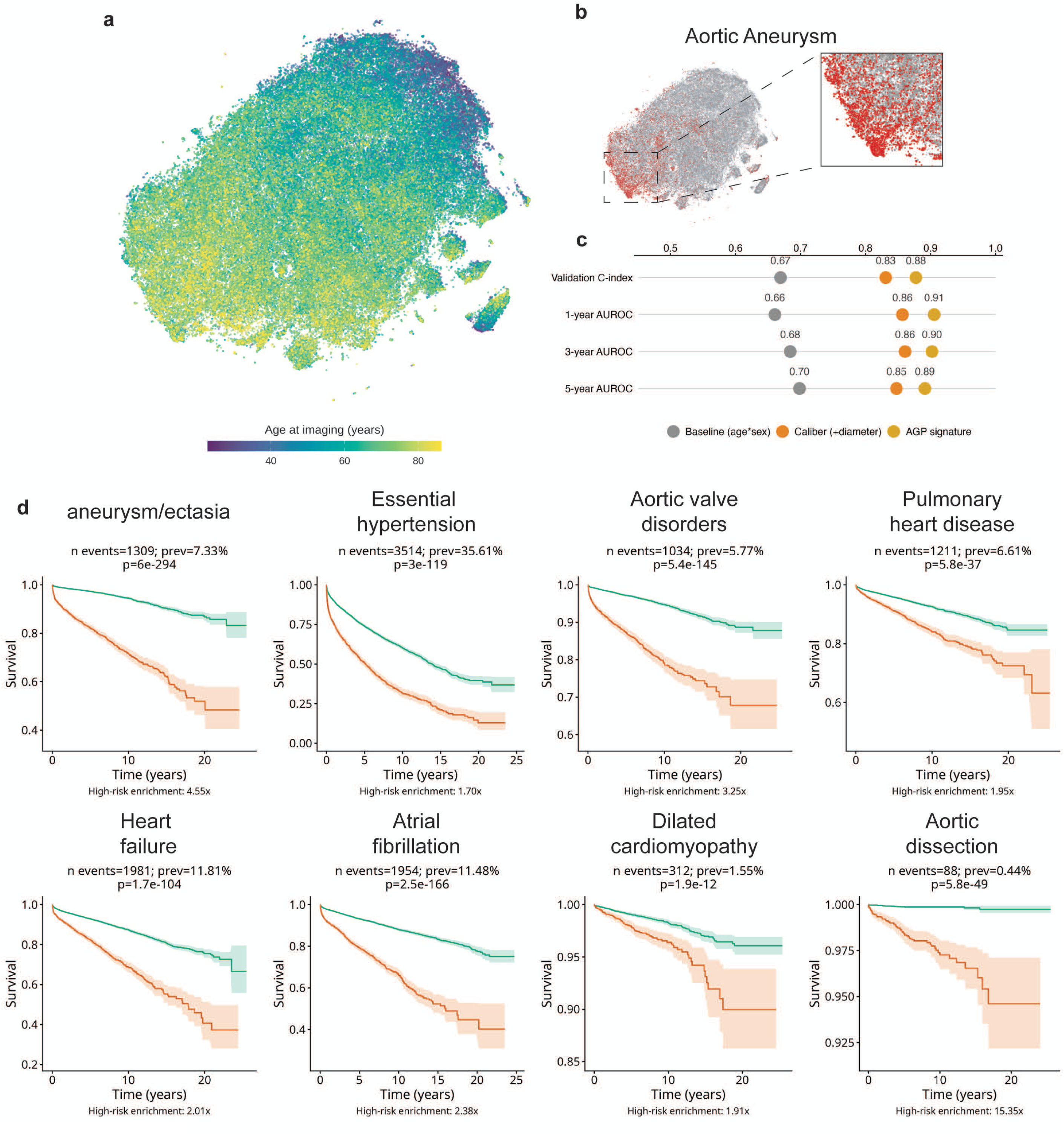
Disease-specific aortic geometric phenotype (AGP) signatures and cardiovascular risk stratification. (**a**) T-distributed stochastic neighbors (t-SNE) embedding of the 38-dimensional AGP space across all participants, colored by age at imaging, demonstrating that the principal axes of geometric variation align with age-related aortic remodeling. (**b**) Aortic aneurysm t-SNE overlay showing focal clustering in a distinct geometric neighborhood. (**c**) Nested model comparison of the baseline (age + sex + age-sex interaction), caliber (baseline + 4 regional aortic diameter measurements), and the AGP signature (baseline + reverse stepwise-selected AGPs) evaluated by validation C-index and IPCW time-dependent AUROC at 1-, 3-, and 5-years. (**d**) Kaplan-Meier curves with 95% confidence intervals for 8 representative cardiovascular diseases in the held-out validation set, stratified into high-and low-risk groups by AGP signature-predicted risk. The risk threshold was set at the 80th percentile of the AGP signature score in the training set and applied to the validation set. Diseases span aortic, valvular, hypertensive, arrhythmic, pulmonary, cardiomyopathy, and heart failure categories. Event counts, prevalence, log-rank P-values, and high-risk enrichment ratios are annotated for each condition.

## DISCUSSION

We present the Aortic Geometric Atlas, a path toward comprehensive, population-scale characterization of thoracic aortic structure across the adult lifespan. By applying the AGT to 62,366 participants, we establish sex-specific aging charts across ages 18 to 99 for a comprehensive set of AGPs, capturing the full geometry of the aorta rather than diameter alone. Several principal findings emerge from this analysis. First, the thoracic aorta undergoes non-linear, phenotype-specific remodeling across the adult lifespan, with distinct trajectories for caliber, length, and shape. Second, non-caliber geometry, including length, tortuosity, curvature, and branch vessel architecture, carries diameter-independent prognostic value for incident disease across the phenome. Finally, disease-specific AGP signatures improve discrimination over caliber-only models, identifying high-risk individuals across diverse cardiovascular conditions. Together, these findings position aortic geometry as a subclinical marker of incident cardiovascular disease and a first step toward a complete atlas of aortic structure.

Prior characterizations of thoracic aortic geometric aging established important foundations in defining its age-related trajectories. The closest prior study to ours, also from our group, examined the geometric aging of the thoracic aorta in the UK Biobank using a deep learning segmentation pipeline followed by multivariate linear regression^11^. While similar in phenotyping strategy, the previous work relied on geometric apex-based landmarks for subsegmental delineation rather than anatomical branch vessel landmarks. Additionally, this method used linear models that cannot capture the non-linear age trajectories observed for many AGPs, nor did it provide centile-based references applicable to individual participants. Earlier studies have similarly been constrained in scope. Turkbey et al.^26^, in the Multi-Ethnic Study of Atherosclerosis (MESA), characterized ascending aortic diameter in 3,573 participants spanning four racial/ethnic groups, but the analysis was limited to a few phenotypes, restricted to ages 45-84, and based on linear regression. Wolak et al.^27^ established age- and sex-stratified diameter limits in 4,039 asymptomatic adults but measured only ascending and descending diameter at a single anatomic level, acquired manually, carrying the overestimation bias noted earlier. Pham et al.^28^ expanded coverage in the Copenhagen General Population Study (n=902) to seven anatomic levels spanning the thoracic and abdominal aorta but similarly relied on manual diameter measurements, providing no broader geometric phenotypes. Adriaans et al.^36^ went beyond diameter to examine aortic length in 210 participants, qualitatively describing the distalward shift of the aortic apex relative to the branch vessels with age. We now quantify this phenomenon through branch vessel takeoff angle trajectories across a substantially larger population. Our framework extends prior studies by providing per-participant centile contextualization of aortic measurements and by characterizing the geometric signatures of distinct aortic pathologies. The AGT codebase is released as an open-source resource to enable future investigations and to standardize geometric phenotyping across institutions.

Beyond these advances, the AAC framework offers immediate clinical utility by transforming aortic assessment from a categorical, threshold-based exercise into a continuous, individualized framework. Clinically reported aortic diameters are typically interpreted against fixed cutoffs (e.g., 4.0 cm for dilation, 5.5 cm for surgical intervention), with body-size indexing for outliers but no continuous referencing to non-linear age- and sex-specific remodeling trajectories observed across the adult lifespan^43^. By contrast, centile-based scoring allows clinicians to identify participants whose aortic geometry deviates meaningfully from age- and sex-matched references even when absolute measurements remain within conventional ranges, potentially enabling earlier identification of accelerated aortic remodeling and more individualized surveillance strategies. Applied to documented pathology, centile scoring resolved condition-specific signatures of increasing complexity, from predominantly caliber-centered shifts in atherosclerosis to multi-phenotype distortion in dissection (**Fig. 4**). Beyond caliber, the dominant non-caliber signal centered on centerline length and descending tortuosity, indicating that pathological remodeling extends past dilation alone. These tortuosity findings are concordant with prior smaller-scale work showing that descending thoracic tortuosity is more pronounced in both aneurysmal and dissected aortas than in unaffected individuals^44^, supporting the framework’s ability to capture clinically meaningful geometry beyond diameter.

Prior work has established that non-caliber measurements of aortic geometry hold independent utility for predicting aortic disease. Heuts et al. identified ascending thoracic aortic length and aortic volume as independent predictors of aortic dissection^15,19^. Wu et al. demonstrated diagnostic utility of ascending aortic length for aortic aneurysm^13^ and Belvroy et al. found descending aortic tortuosity to be associated with both aneurysm and dissection^44^. Our phenome-wide time-to-event analysis confirms and extends these findings at population scale: aortic aneurysm was associated with 21 AGPs spanning 7 phenotype categories, with 11 associations involving non-caliber phenotypes including centerline length, tortuosity, and taper ratio, each carrying independent prognostic value beyond regional diameter measurements. The association of aortic centerline length with incident atrial fibrillation further extends the prognostic reach of non-caliber phenotypes beyond aortic disease, consistent with prior work linking aortic elongation and arch remodeling to adverse left ventricular remodeling^37^. Heart failure showed a comparable pattern across subtypes, with associations somewhat stronger in HFpEF, consistent with ventricular-arterial coupling in diastolic dysfunction^39–42^. Beyond individual associations, combining multiple AGPs into disease-specific signatures improved prediction of incident cardiovascular disease over caliber-based and baseline models, indicating that clinically actionable information is distributed across many phenotypes rather than concentrated in diameter. This is consequential because diameter anchors current risk assessment yet captures only part of the relevant geometry, as aortic dissections commonly occur below the diameter cut points outlined in clinical guidelines^45^. By integrating non-caliber features such as length, tortuosity, and branch vessel architecture, AGP signatures begin to supply this additional information, with the largest gains where structural remodeling is most pronounced, as in aortic aneurysm. In aggregate, this supports AGPs as useful subclinical markers of cardiovascular disease, with the potential to flag risk that emerges before, or independent of, changes in diameter.

Several considerations should be noted when interpreting these findings. Both cohorts comprise clinically indicated CT studies rather than population-based screening, so these analyses are subject to index event bias, whereby selection on the clinical indication for imaging can distort observed associations. Reference participants were defined by the absence of documented aortic disease rather than by health screening, so they may include individuals with subclinical disease. The phenome-wide time-to-event analysis is retrospective, and residual confounding cannot be excluded. Third-order phenotypes such as eccentricity and torsion showed the lowest reproducibility, limiting their clinical utility, since their reliable estimation exceeds what current clinical voxel spacing provides. Additionally, the geometric profiles observed in participants with documented aortic pathology (e.g., dissection, coarctation) may reflect the effects of prior surgical or endovascular intervention rather than untreated disease alone. Future work should prioritize prospective validation of the AAC framework for clinical decision-making, including surveillance planning, external validation across geographically and demographically diverse populations, and evaluation using high-resolution ECG-gated CT acquisitions to improve reliability of higher-order geometric phenotypes.

In summary, the Aortic Geometric Atlas establishes a comprehensive, population-scale reference framework for thoracic aortic geometry across the adult lifespan. The accompanying phenome-wide time-to-event analysis demonstrates that aortic geometry is prognostic across a wide range of incident conditions, with non-caliber phenotypes contributing predictive value that diameter alone does not capture. Together, these resources mark an initial step toward a complete structural characterization of the aorta.

## METHODS

### Penn Medicine BioBank imaging data

The three-dimensional computed tomographic imaging data used in this study were from the Penn Medicine BioBank (PMBB). The PMBB has enrolled more than 350,000 consenting participants who receive care in the Penn Medicine healthcare system^30^. Participants were enrolled during routine clinical care appointments at a Penn Medicine location during which they provided written consent either in-person or digitally to participate in the PMBB. This consent enabled the linkage of their electronic health records, including past disease diagnosis, laboratory measurements, and imaging data. The PMBB was approved by the Institutional Review Board at the University of Pennsylvania under IRB protocol 813913. The PMBB data is available under a data use agreement with the University of Pennsylvania, with access governed by HIPAA-compliant restrictions on re-identification, external sharing, and commercial use. This research was approved by the Institutional Review Board at the University of Pennsylvania under IRB protocol 857974.

### CT-RATE dataset

The CT-RATE dataset includes 50,188 CT volumes and corresponding radiology reports acquired from 25,692 studies performed on 21,304 participants (ages 18-102, 57.5% male) at the Istanbul Medipol University Mega Hospital between 2015 and 2023^31^. The participants’ age at the time of imaging was reported as an integer rounded to the nearest year. After quality control and exclusion of studies without TotalSegmentator segmentations, 50,171 CT volumes across 25,685 studies from 21,299 participants were retained for analysis. Additional details regarding imaging metadata, postprocessing, and quality control are available at: https://huggingface.co/datasets/ibrahimhamamci/CT-RATE.

### Reference cohort definition

To establish reference ranges for aortic geometry, we identified participants without evidence of aortic disease from both cohorts. Aortic findings were first extracted from radiology reports using a large language model (GLM-4^46^), which parsed each report into a structured template of 15 aortic conditions including aneurysm, ectasia, atherosclerosis, dissection, intramural hematoma, mural thrombus, pseudoaneurysm, rupture, stenosis, coarctation, aortitis, penetrating ulcer, infected aneurysm, primary tumor, and postsurgical changes. Any study with a documented aortic finding was excluded from the reference cohort. For the PMBB cohort, additional exclusions were applied using ICD-10 diagnosis codes drawn from each participant’s complete clinical history, regardless of timing relative to the imaging date. Participants were excluded if they had any prior diagnosis of congenital aortic anomalies, bicuspid aortic valve, aortic aneurysm or dissection, connective tissue disorders associated with aortopathy, large vessel vasculitis, prior aortic or cardiac surgery, atherosclerosis of the aorta, or secondary causes of aortic remodeling (**Table S13**). For the CT-RATE dataset, exclusions were based solely on radiological report findings.

### Aortic Geometric Toolkit (AGT) overview

We developed a fully automated pipeline for extracting geometric phenotypes of the thoracic aorta from computed tomography (CT) segmentations produced by TotalSegmentator^1^. The pipeline accepts a multi-label segmentation volume containing the aorta (label 52) and thoracic vertebrae, extracts a binary thoracic aorta mask cropped at the T12 vertebral level, and computes a comprehensive set of morphological measurements including diameter, length, curvature, torsion, tortuosity, cross-sectional area, eccentricity, volume, and branch vessel geometry. AGPs are organized by the order of derivation from the underlying centerline. **First-order** phenotypes are direct geometric measurements, including length, diameter, volume, surface area, cross-sectional area, arch width, and arch height. **Second-order** phenotypes are derived through differentiation or ratios of first-order quantities and include curvature, tortuosity, and taper ratio. **Third-order** phenotypes capture local cross-sectional shape or higher-order centerline geometry and include eccentricity and torsion. Complete definitions of all extracted phenotypes and the TotalSegmentator label identifiers used in the pipeline are provided in **Tables S14** and **S15**, respectively. All processing was implemented in Python using open-source libraries (scikit-image, scipy, numpy, SimpleITK). Complete AGT code is available at https://github.com/cams2b/aortic_geometry_toolkit.

### Thoracic aorta extraction and skeletonization

The aorta label (ID 52) was extracted from the TotalSegmentator multi-label segmentation as a binary mask^29^. To restrict the analysis to the thoracic aorta, voxels inferior to the T12 vertebra (label 32) were zeroed, with the cropping direction determined by comparing the median z-slice indices of T1 (label 43) and T12 to account for scan orientation. The binary mask was cleaned via morphological closing with a spherical structuring element (radius 2 voxels) to fill small gaps, followed by per-slice hole filling, extraction of the largest connected component, and trimming of empty boundary slices. The cleaned mask was skeletonized to a 1-voxel-wide medial axis using a 3D parallel thinning algorithm that preserves topological connectivity^47^. For scans with any axis spacing exceeding 1.5 mm, the mask was resampled to 1 mm isotropic resolution prior to skeletonization to ensure adequate resolution of tortuous regions. Empty z-slices at the superior and inferior boundaries were trimmed, and the image origin was adjusted to maintain correct physical coordinates.

### Centerline extraction

Prior to skeletonization, the Euclidean distance transform (EDT) was computed on the binary mask with anisotropic voxel spacing. At each skeleton voxel, the EDT value represents the radius of the maximal inscribed sphere, the distance to the nearest background voxel, providing the vessel radius at each centerline point:

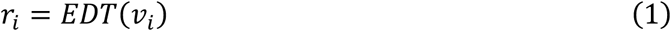

where *v_i_* is the skeleton voxel at centerline point *i*. The diameter at each point is *d_i_* = 2*r_i_*. Next, a voxel adjacency graph was constructed from the skeleton using 26-connectivity, where each skeleton voxel constitutes a node and edges connect voxels within a 3×3×3 neighborhood. Skeleton endpoints (degree-1 nodes) and junction points (degree ≥3 nodes, representing aortic branch take-off sites) were identified from the graph structure. The main aorta trunk was extracted as the longest path between any two endpoints using the standard tree-diameter algorithm^48^: two-pass breadth-first search (BFS) with physical distance weighting. The first pass from an arbitrary endpoint identifies one extreme of the tree; the second pass from that extreme identifies the true farthest endpoint and records the predecessor chain for path reconstruction. This approach intrinsically excludes branch vessel spurs because the main ascending, arch, and descending path is always the longest path in the skeleton graph. The ordered path was smoothed with a one-dimensional Gaussian kernel applied independently to each spatial coordinate to reduce voxel-level staircase artifacts, then fitted with a cubic B-spline parameterized by cumulative chord length and resampled to N = 200 uniformly spaced points. Distance-transform radii were interpolated onto the resampled centerline via linear interpolation, yielding a final representation of N points each with physical coordinates (x, y, z) and an associated vessel radius in millimeters.

### Anatomical region definition

When the brachiocephalic trunk (TotalSegmentator label 54) and left subclavian artery (label 56) were present in the segmentation, the aortic arch was defined anatomically between the two branch take-off points along the centerline. Branch junction coordinates were identified by dilating the aorta mask by 2 voxels, computing the intersection with each branch label, and taking the centroid of the resulting junction zone. The closest centerline points to each junction coordinate were used as region boundaries, partitioning the centerline into the ascending aorta (aortic root to brachiocephalic trunk origin), aortic arch (brachiocephalic trunk to left subclavian artery origin), and descending aorta (left subclavian artery to the T12 vertebral level). When branch landmarks were unavailable, regions were instead defined by the z-coordinate (superior-inferior) profile of the centerline: the arch apex was identified as the point of maximum z, the arch region as the portion of the centerline exceeding 70% of the total z-range, and the ascending and descending segments as the portions proximal and distal to the apex, respectively.

### Geometric phenotypes

The following measurements were computed for each anatomical region (overall, ascending aorta, aortic arch, descending aorta) unless otherwise noted.

#### Centerline Length

The arc length of each region was computed as the sum of Euclidean distances between consecutive centerline points:

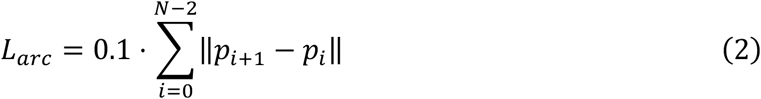

where *p_i_* is the *i*-th centerline point in physical (mm) coordinates and the factor 0.1 converts mm to cm.

#### Diameter

Diameter was derived from the distance-transform radii at each centerline point (Equation 1). Per-region summary statistics (mean, median, minimum, maximum, standard deviation) were computed from the diameter profile in centimeters.

#### Tortuosity

Tortuosity was defined as the ratio of arc length to chord length minus one:

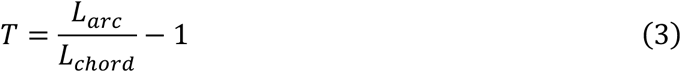

where *L_arc_* is the centerline arc length (Equation 2) and *L_chord_* is the Euclidean distance between the first and last centerline points of the region. A value of 0 indicates a perfectly straight vessel; higher values indicate greater tortuosity.

#### Curvature

Curvature was computed at each centerline point using the Frenet-Serret formulation. First and second spatial derivatives of the centerline were estimated using central finite differences via numpy’s gradient function. Curvature κ at each point was computed as:

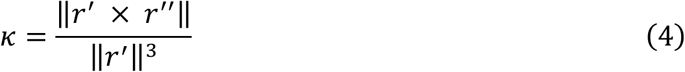

where *r*′ and *r*″ are the first and second derivatives of the centerline and × denotes the cross product. The first and last 3 points were excluded from summary statistics to avoid edge effects. Mean and maximum curvature were reported per region.

#### Torsion

Torsion measures the rate at which the curve twists out of its local osculating plane:

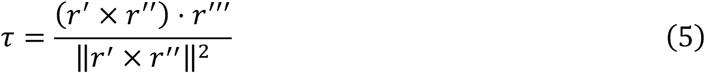

where *r′″* is the third derivative and ⋅ denotes the dot product. A planar curve has zero torsion. The same boundary trimming was applied as for curvature. Mean torsion was reported per region. Torsion was retained in the AGT pipeline as a standard differential geometric property of three-dimensional curves but was excluded from downstream centile analyses due to high sensitivity of the discrete third-derivative computation to centerline noise on non-gated CT, which produced poor test-retest reproducibility (**Supplemental Materials**).

#### Cross-sectional area

Cross-sectional area was estimated at each centerline point from the distance-transform radius, assuming a circular cross-section:

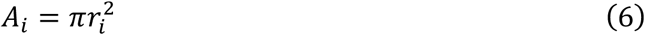

The mean cross-sectional area was reported per region in cm².

#### Eccentricity

Cross-sectional eccentricity was estimated at sampled centerline points (approximately 20 per region) to quantify deviation from circularity. At each sampled point, aorta mask voxels within 30 mm were identified and projected onto the plane perpendicular to the local centerline tangent. Voxels within 2 mm of the plane were retained, and their radial distances from the centerline were computed. Eccentricity was defined as:

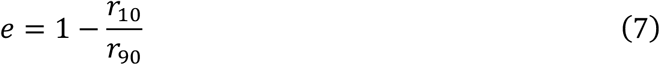

where *r*_10_ and *r*_90_ are the 10th and 90th percentile radii, respectively. Values range from 0 (circular) to 1 (maximally elongated). Mean eccentricity was reported for the overall, ascending, and descending regions.

#### Taper ratio

The taper ratio quantifies the change in vessel caliber along a region:

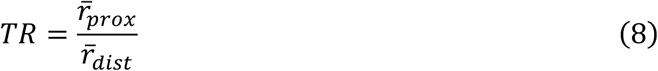

where *r̅_prox_* and *r̅_dist_* are the mean radii of the first and last 10% of the region’s centerline points. Values greater than 1 indicate distal narrowing.

#### Arch height and width

Arch height was defined as the vertical (superior-inferior) extent of the aortic arch centerline:

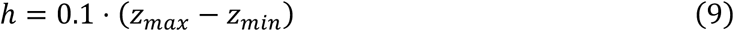

Arch width was the Euclidean distance between the arch endpoints:

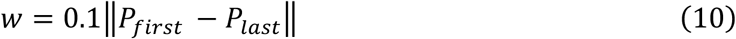

#### Volume and surface area

Aortic volume was computed by voxel counting:

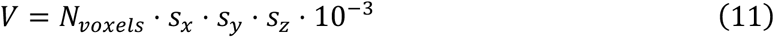

where *N_voxels_* is the number of foreground voxels and *s_x_*, *s_y_*, *s_z_* are the voxel spacings in mm. Surface area was computed from a smoothed triangle mesh generated by the marching cubes algorithm^49^ with 200 iterations of Taubin^50^ smoothing applied to reduce staircase artifacts:

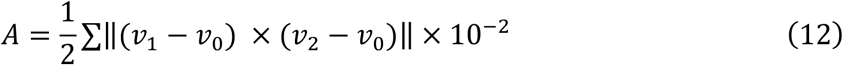

where the summation is over all triangles in the mesh, *v*_0_, *v*_1_, and *v*_2_ are the vertex coordinates of each triangle in mm, and × denotes the vector cross product whose magnitude equals twice the triangle area.

#### Branch take-off angles

For each detected branch vessel (brachiocephalic trunk, left subclavian artery, left common carotid artery), the take-off angle relative to the local aorta direction was computed. The local aorta tangent was estimated from centerline points ±2 positions from the branch junction. The branch direction vector was defined from the junction point toward the centroid of all branch vessel voxels. The angle was computed as:

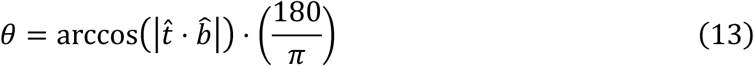

where *t̂* is the unit tangent, *b̂* is the unit branch direction, ⋅ denotes the dot product, and the factor 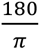 converts radians to degrees. The absolute value ensures the angle is reported in [0°, 90°].

### Aortic Aging Charts construction

We modeled age-related trajectories in aortic geometric phenotypes using generalized additive models for location, scale, and shape (GAMLSS)^32,33^. Following previous work^34,35^, for each phenotype we evaluated three candidate distribution families: the Box-Cox Cole-Green (BCCGo) distribution, which models the mean, scale, and skewness as functions of age; the Box-Cox t (BCTo) distribution, which additionally accommodates heavy-tailed data via a kurtosis parameter; and the normal (NO) distribution as a two-parameter baseline. For each phenotype, the response variable y was assumed to follow the selected distribution:

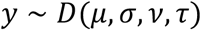

where μ (location) and σ (scale) were modeled as functions of age using fractional polynomials with sex as an interaction term, while ν (skewness) and τ (kurtosis), when present, were held constant (intercept-only). We defined a grid of three complexity levels for the pair (μ order, σ order): (1,1), (2,1), and (2,2), yielding 9 candidate model specifications per phenotype (3 fractional polynomial configurations × 3 distribution families). Outliers beyond 3.5 standard deviations from the sex-specific mean were excluded prior to model fitting. The best fitting model was selected by the minimization of the Bayesian information criterion (BIC). An example model for a BCCGo distribution with second-order fractional polynomials on *μ* and first-order fractional polynomials on *σ* (fp(2,1)), the model takes the form:

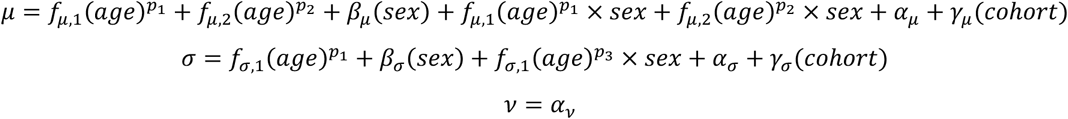

where the f terms describe the fixed effects of age, with the fractional polynomial powers optimized from the candidate set [−2, −1, −0.5, 0, 0.5, 1, 2, 3], the sex interaction terms enable sex-specific trajectories in both the expected value and dispersion of each phenotype, and γ(cohort) denotes a random intercept accounting for systematic differences across acquisition protocols and imaging sites. We evaluated two random effect specifications: cohort alone (PMBB vs CT-RATE) and scanner x cohort, where each unique combination of harmonized scanner model and cohort defined a distinct level (74 levels total). Scanner model was determined from the DICOM ManufacturerModelName field and harmonized across cohorts using a mapping function that consolidated duplicate naming conventions, excluded post-processing software entries, and grouped models with fewer than 20 studies, yielding 67 scanner groups in PMBB and 7 in CT-RATE. This approach identifies distinct scanner models rather than individual physical scanners, as unique device identifiers were not available in either dataset; however, it captures variation across scanner generations, including older scanners replaced by newer models at the same site. For the scanner x cohort specification, the convergence tolerance was relaxed from 0.001 to 0.01.

### Sensitivity analysis: bootstrap resampling

To assess uncertainty in the estimated centile and rate-of-change curves, we performed bootstrap resampling with 1,000 iterations using stratified sampling with replacement, preserving both the sex ratio and cohort composition. For each iteration, the best-fitting model specification was refit to the resampled data using the original fit’s predicted values as initialization to improve convergence. Centile curves at the 2.5th, 25th, 50th, 75th, and 97.5th percentiles were recomputed on a fine age grid spanning the observed age range, and rates of geometric change were estimated by numerical differentiation of the fitted centile curves. Empirical 95% confidence intervals (2.5th-97.5th percentiles of the bootstrap distributions) were computed for each age point on the centile and rate-of-change curves. For each phenotype and sex, characteristic ages of interest were extracted, including the ages at which the median centile curve attained its minimum and maximum, the ages of peak positive and peak negative rates of change, and the earliest age at which the rate-of-change crossed zero. Bootstrap confidence intervals for these characteristic ages were computed from the corresponding bootstrap distributions.

### Aortic pathological scoring

The fitted reference GAMLSS models provided the basis for characterization of aortic pathologies. For each study with documented aortic pathology in the radiology report, the cumulative distribution function of the sex- and age-stratified reference was applied to each AGP value to yield an individual-level centile score (0-1) representing the participants’ relative position within the reference distribution. These centiles provide a population- and age-standardized clinical phenotype that enables direct cross-AGP and cross-pathology comparison. Deviations from the population median (centile = 0.5) were taken to reflect pathology-specific geometric signatures. For each AGP and pathology, the magnitude of deviation from the AAC median was quantified using Cohen’s D, computed as the difference between the mean centile and 0.5 divided by the standard deviation of participants centile values. The statistical significance of deviations was assessed with two-sided Wilcoxon signed-rank tests against a null hypothesis of centile = 0.5, with Bonferroni correction applied to identify AGPs that differ significantly from the AAC reference (*P* < 0.05 / (38 AGPs x 4 diseases)).

### Classification of prevalent aortic disease

To establish the generalizability of AGP-based disease classification across institutions, we trained logistic regression classifiers in PMBB and externally validated them in the CT-RATE dataset. Models predicted prevalent aortic disease (aneurysm, atherosclerosis, and dissection), as documented in the radiology report, using the 38 AGPs. AGPs were standardized using the training-set mean and standard deviation (z-score normalization), with the held-out data transformed by these same parameters. Predictors were selected within each training set by reverse stepwise elimination, in which all 38 AGPs were initially included and the AGP with the largest P-value was iteratively removed until all remaining terms achieved nominal significance (*P* < 0.05). Internal performance in PMBB was estimated by repeated grouped cross-validation (5 repeats of 10-fold cross-validation), with folds assigned at the patient level so that all scans from a patient shared a single fold. The AUROC and Brier score were computed on each held-out fold. The mean and 95% confidence interval were taken across the 50 resamples. The final model was fit on the full PMBB cohort and was then applied to CT-RATE, with 95% confidence intervals determined from 1,000 bootstrap resampling.

### Phenome-wide time-to-event analysis

The prognostic value of aortic geometric phenotypes was assessed through a phenome-wide time-to-event analysis for incident disease in the PMBB cohort using a stratified 50-50 discovery-replication design. First, ICD-9/10CM codes were mapped to phecodes using PheCodeX (https://github.com/PheWAS/PhecodeX)^51^. Following the rule-of-two, a phecode qualified as an incident event only if it was recorded on at least two distinct dates, with the first occurrence after the index date^52^. For each phecode with at least 50 incident cases per split, we used the first CT in each participant’s medical record as the index date. Participants with a prevalent diagnosis (recorded at or before the index date) were excluded from the analysis for that phecode. Follow-up was censored at the date of the last recorded clinical encounter in the electronic health record. Cox proportional hazards models were fit at the individual phecode level with the AGP as the primary predictor, adjusting for age at imaging, sex, and an age-sex interaction as covariates^53^. To enable comparison of effect sizes across phenotypes, all AGPs were z-score normalized within each split independently to prevent data leakage. To identify AGPs with prognostic value independent of aortic caliber, we fit a second set of models in which the four regional diameter measurements (ascending, arch, descending, and overall) were included as additional covariates; the incremental discriminative value of each non-diameter AGP was quantified by the change in Harrell’s concordance index (ΔC-index) relative to the diameter-adjusted baseline model. Associations were considered significant if they met Bonferroni correction for the total number of tests performed (*P* < 0.05 / (number of AGPs x number of phecodes)) with concordant hazard ratio direction across both discovery and replication splits.

### t-SNE visualization of aortic geometric phenotypes

The AGP feature space was visualized using T-distributed stochastic neighbor embedding (t-SNE) applied to the z-score standardized 38-dimensional AGP vectors across all patients. t-SNE was computed using the Barnes-Hut approximation (θ=0.5) with perplexity 50, 1,000 iterations, and an initial projection to 30 principal components, with age and disease labels overlaid to assess geometric clustering patterns.

### Disease-specific AGP signatures

To evaluate the combined prognostic value of aortic geometric phenotypes for incident cardiovascular risk prediction, we developed disease-specific geometric signatures using multivariable Cox proportional hazards models. For each cardiovascular phecode with at least 50 incident cases in the PMBB cohort, we constructed three nested Cox models. The baseline model included age at imaging, sex, and an age-sex interaction. The caliber model added subsegmental diameter measurements of the ascending aorta, aortic arch, descending aorta, and overall aortic diameter to the baseline. The AGP signature model incorporated geometric phenotypes selected via reverse stepwise elimination. For the signature model, all 38 standardized AGPs were initially included alongside baseline covariates, and the AGP with the largest P-value was iteratively removed until all remaining AGP terms achieved nominal significance (*P* < 0.05). Baseline covariates were retained regardless of significance.

Model evaluation followed a stratified 50-50 discovery-validation design analogous to the phenome-wide time-to-event analysis. For each disease, the cohort was split 50/50 into training and validation sets, stratified on case status. AGPs were standardized using the training-set mean and standard deviation (z-score normalization), with the validation set transformed by these same parameters. Prevalent cases observed before the time of imaging were excluded and follow-up was censored at the date of last recorded clinical encounter. Model discrimination on the held-out validation split was assessed using Harrell’s C-index alongside the time-dependent AUROC and Brier score. Both were evaluated at 1-, 3-, and 5-years under inverse probability of censoring weighting (IPCW), as implemented in the riskRegression R package (https://github.com/tagteam/riskRegression). For each metric, 95% confidence intervals were computed using 1,000 bootstrap resamples of the validation split, stratified by event status. The incremental discrimination of the AGP-signature over the baseline model, as well as the caliber model, was tested by paired comparison of time-dependent AUROC using the method of Blanche et al., a censored-data generalization of DeLong’s test^54,55^. To account for multiple comparisons, a 5% FDR correction was applied across the 5-year time-dependent AUROC comparisons, with gains surviving correction considered significant. Risk stratification used a threshold set at the 80th percentile of the training-split linear predictor, applied to the validation split to define high-risk (top 20%) versus low-risk (bottom 80%) groups. Separation between groups was assessed by the log-rank test with Bonferroni correction. Enrichment was calculated as the ratio of observed event rates between high- and low-risk groups.

## Supporting information

Supplemental Note: PMBB leadership

## CODE AVAILABILITY

Aortic geometric toolkit (AGT) code is available on GitHub: https://github.com/cams2b/aortic_geometry_toolkit

## DATA AVAILABILITY

Access to Penn Medicine BioBank (PMBB) data is provided to investigators at the University of Pennsylvania. Individual-level data from the Penn Medicine BioBank (PMBB) are subject to institutional data use agreements and participants privacy regulations, and access is limited to qualified researchers via the PMBB application process: https://pmbb.med.upenn.edu/investigators.php. CT-RATE data is available on Hugging Face: https://huggingface.co/datasets/ibrahimhamamci/CT-RATE.

## ACKNOWLEDGEMENTS

We acknowledge the Penn Medicine BioBank (PMBB) for providing data and thank the patient- participants of Penn Medicine who consented to participate in this research program. The PMBB is approved under IRB protocol# 813913 and supported by Perelman School of Medicine at University of Pennsylvania, a gift from the Smilow family, and the National Center for Advancing Translational Sciences of the National Institutes of Health under CTSA award number UL1TR001878. Research reported in this publication was supported by the National Heart, Lung, And Blood Institute of the National Institutes of Health under Award Number F31HL182332. The content is solely the responsibility of the authors and does not necessarily represent the official views of the National Institutes of Health.

## FUNDING

C.A.B is supported by NIH grant F31-HL182332. W.R.W. is supported by NIH grants P41-EB029460, R01-HL169378, R01-HL137984, UL1-TR001878, R21-EB036734, OT2-OD038048. J.A.C. is supported by NIH grants R01-HL157108, U01-HL160277, and K24-AG070459. J.G is supported by R01-EB031722, R01-HL133889. S.Z. is supported by NIH grant T32-DK007740.

## DISCLOSURE

J.A.C is supported by NIH grants U01-TR003734, U01-TR003734-01S1, UO1-HL160277, R33-HL-146390, R01-HL153646, K24-AG070459, R01-AG058969, R01-HL157108, R01-HL155599, R01-HL104106 and R01HL155764. He has recently consulted for Bayer, Fukuda-Denshi, Bristol-Myers Squibb, Biohaven Pharmaceuticals, Johnson & Johnson, Edwards Life Sciences, Merck, NGM Biopharmaceuticals, S2N Health, Health Advances, Emory University and University of Delaware. He received University of Pennsylvania research grants from National Institutes of Health, Fukuda-Denshi, Bristol-Myers Squibb, Microsoft and Abbott. He is named as inventor in a University of Pennsylvania patent for the use of inorganic nitrates/nitrites for the treatment of Heart Failure and Preserved Ejection Fraction and for the use of biomarkers in heart failure with preserved ejection fraction. He has received payments for editorial roles from the American Heart Association, the American College of Cardiology, Elsevier and Wiley, and payments for academic roles from the University of Texas, Boston University, Rochester Regional Health, Virginia Commonwealth University and the Korean Vascular Society. He has received research device loans from Atcor Medical, Fukuda-Denshi, Unex, Uscom, NDD Medical Technologies, Microsoft and MicroVision Medical.

## Notes

### Author Declarations

This research was approved by the Institutional Review Board at the University of Pennsylvania under IRB protocol 857974.

