## Supplemental Material for "Aortic Geometric Atlas: Centile-Based Reference Charts and Pathological Signatures Across the Adult Lifespan"

***Corresponding author:**

Cameron A. Beeche, BS

South Tower, Rm. 11-155.

**SUPPLEMENTAL EXTENDED RESULTS**

**50-50 discovery-replication sensitivity analysis**

To assess the stability of the aging charts against random sampling variation, we performed a stratified 50-50 discovery-replication split of the reference cohort, stratified by sex and age group (18-30, 30-40, 40-50, 50-60, 60-70, 70-80, 80-90, ≥90 years) to preserve the joint age-sex distribution. Each half was refit independently, performing automated model selection across the same 9 candidate specifications. Median centile curves and rate-of-change curves from the two halves were then compared across the full adult age range for each sex. Agreement was quantified as the Pearson correlation (R), coefficient of determination (R2), mean absolute difference (MAD), and root mean square error (RMSE) between the two curves. Across the 38 AGPs, centile curves showed high agreement in both sexes (mean R²=0.990 male, 0.983 female), with R² above 0.95 in 37 of 38 phenotypes for males and 35 of 38 for females. Rate-of-change curves were similarly consistent (mean R²=0.922 male, 0.926 female), with R² above 0.95 in 29 of 38 and 32 of 38, respectively. The lowest agreement was observed among higher-order phenotypes (taper ratio, curvature, and torsion). Per-AGP agreement metrics are reported in **Table S4**, and representative discovery-replication overlays are shown in **Figs. S9-S10**.

**Scanner random effect sensitivity analysis**

To assess the sensitivity of the aging charts to scanner-level variation, we refit the full reference cohort using an alternative random effect specification in which the cohort-level random intercept was replaced by a scanner x cohort random effect, such that the same scanner model appearing in both PMBB and CT-RATE was treated as two distinct random effect levels (74 levels total). Automated model selection across the same 9 candidate specifications was performed independently for the scanner-crossed models, and the convergence tolerance was relaxed from 0.001 to 0.01 to accommodate the increased number of random effect levels. Median centile curves and rate-of-change curves from each specification were then compared across the full adult age range for each sex. Agreement was quantified using the Pearson correlation (R), coefficient of determination (R2), mean absolute difference (MAD), and root mean square error (RMSE). Across the 38 AGPs, centile curves showed high agreement between the cohort and scanner-crossed specifications (mean R²=0.972 male, 0.982 female), with R² above 0.95 in 35 of 38 phenotypes for males and 36 of 38 for females. Rate-of-change curves showed greater sensitivity to random effect specification (mean R²=0.954 male, 0.939 female), with R² above 0.95 in 31 of 38 and 30 of 38, respectively. The lowest centile agreement was observed for higher-order phenotypes, including ascending eccentricity and ascending tortuosity. Per-AGP agreement metrics are reported in **Table S5**, and centile and rate R^2^ scatter plots are shown in **Figs. S11-S12**.

**Longitudinal centile stability**

Quantitative geometric assessment of the aorta on non-gated CT is intrinsically affected by the cardiac cycle, since the aorta is a pulsatile vessel whose diameter, length, and curvature change with each ventricular contraction^1^. Without ECG gating, each scan captures the aorta at an essentially random phase of the cardiac cycle, and this physiological pulsatility constitutes an irreducible source of measurement variability between repeated scans of the same participant. With this limitation in mind, we sought to assess longitudinal reproducibility of AAC centile assignments by identifying subjects in PMBB and CT-RATE with two or more CT studies passing AGT processing. For each AGP, we scored every available scan against the full-cohort GAMLSS fit using the participant's age and sex at the time of the scan, yielding a centile value (0–1) per study. For each participant, scans were sorted in ascending order by age and consecutive pairs were constructed, with the interval defined as the difference in age (in years) between paired studies. For each pair we computed the absolute centile difference between the two studies. Per-AGP stability metrics included the median absolute centile difference, the interquartile range, the proportion of pairs with absolute centile difference exceeding 0.05 and 0.10, and the Pearson correlation between paired centile values. Metrics were computed globally and stratified both by cohort (PMBB versus CT-RATE) and by interval bin (<0.1, 0.1–0.5, 0.5–1, 1–3, and >3 years between paired studies).

**Longitudinal stability among first order phenotypes**

Across 23,919 paired studies from participants in PMBB and CT-RATE with two or more CT examinations, the median absolute centile difference between consecutive scans, taken across all 38 AGPs, was 0.114 (range: 0.054, 0.245), with a median Pearson correlation of 0.67 between paired centile values (range: 0.07-0.89). Stability tracked the order of derivation from the underlying centerline. Among first-order phenotypes, global measurements that integrate geometric information across the entire aorta were the most reproducible, with total aortic volume showing the highest test-retest stability of any AGP (R=0.89, median absolute centile difference 0.054; Interquartile range (IQR)=0.020-0.117; **Fig. S13**) followed by surface area (R=0.88, median=0.057; IQR=0.021-0.123; **Fig. S14**), overall cross-sectional area (R=0.80, median=0.086; IQR=0.032-0.183; **Fig. S15**), overall diameter (R=0.78, median=0.089; IQR=0.034-0.190; **Fig. S16**), and overall centerline length (R=0.75, median=0.098; IQR=0.038-0.205; **Fig. S17**). Among regional first-order phenotypes, a clear proximal-to-distal gradient was observed for both diameter and length. Ascending aortic diameter (R=0.70, median absolute centile difference 0.115; IQR=0.045-0.235) and arch diameter (R=0.66, median=0.106; IQR=0.038-0.236) were less reproducible than descending aortic diameter (R=0.79, median=0.084; IQR=0.032-0.181; **Fig. S18**). Similarly, ascending aortic length (R=0.56, median=0.155; IQR=0.059-0.313) was less reproducible than descending aortic length (R=0.75, median=0.087; IQR=0.033-0.190) (**Fig. S19**). This pattern is biologically consistent with regional differences in aortic wall composition. The ascending aorta is elastin-rich and highly distensible, undergoing greater pulsatile distension and motion with each cardiac cycle, whereas the descending aorta is collagen-rich and more mechanically constrained by surrounding structures including the spine and intercostal arteries. Phenotypes measured in the more compliant proximal segments are therefore more affected by cycle-phase variability on non-gated CT than those measured distally.

**Longitudinal stability among higher-order phenotypes**

Second-order phenotypes derived through differentiation or ratios of first-order quantities showed substantially greater variability (median R=0.42), with curvature (**Fig. S20**), tortuosity (**Fig. S21**), and taper ratio (**Fig. S22**) all showing reduced reproducibility relative to first-order measurements. Descending aortic tortuosity retained the highest reproducibility among all second-order phenotypes (R=0.83, median absolute centile difference: 0.081), approaching the stability observed for integrated first-order measurements. A consistent proximal-to-distal gradient was observed across all second-order categories, with ascending aortic tortuosity, curvature, and aortic taper ratio all showing markedly reduced reproducibility compared to their descending counterparts, mirroring the pattern observed among first-order regional phenotypes and again consistent with greater cardiac-cycle-dependent variability in the more compliant, elastin-rich proximal aorta. Third-order phenotypes capturing local cross-sectional shape or higher-order centerline geometry (torsion and eccentricity) showed the lowest reproducibility of all (median R=0.20, range=0.07-0.24) (**Figs. S23**-**S24**). This pattern is consistent with the cardiac-cycle dependence of aortic geometry on non-gated CT^1^. The aorta is a pulsatile vessel whose luminal diameter, length, and curvature change with each ventricular contraction, with reported ascending aortic diameter changes between systole and diastole ranging from approximately 3% to over 20% across studies^2,3^, and substantial longitudinal pulsatile changes also reported in the proximal aorta^4^. Without ECG gating, each scan captures the aorta at an essentially random phase of the cardiac cycle, and this physiological pulsatility constitutes an irreducible source of measurement variability in standard clinical CT imaging. Higher-order phenotypes additionally rely upon a robust underlying centerline for their computation, the quality of which has a lower bound set by image resolution, cardiac motion, participant motion, and segmentation accuracy. As phenotypes move further from the underlying centerline through successive abstractions from first-order to second-order to third-order quantities, small perturbations in centerline placement are progressively amplified, increasing measurement noise at each level of derivation. Integrated first-order phenotypes, by contrast, average over both cycle-dependent local variations and centerline perturbations and are therefore most robust. Per-AGP longitudinal stability metrics for all 38 AGPs are provided in **Table S7**.

**SUPPLEMENTAL FIGURES**

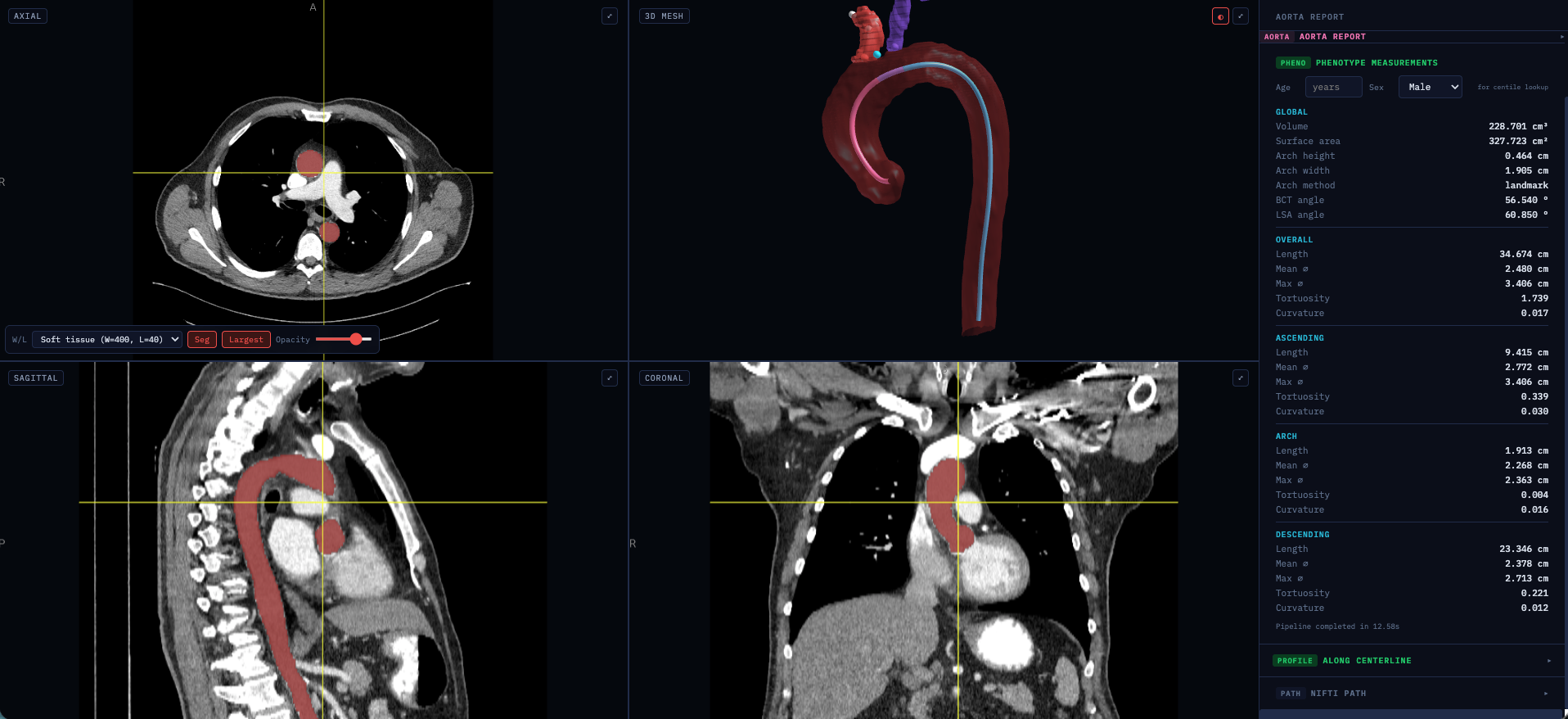

**Figure S1. Aortic Geometry Toolkit (AGT) interactive visualizer.** Screenshot of the AGT web-based visualizer demonstrating the complete geometric phenotyping output for a representative participant with a contrast-enhanced CT volume. The interface displays synchronized axial (top-left), sagittal (bottom-left), and coronal (bottom-right) CT views with aortic segmentation overlay, a three-dimensional surface mesh with anatomical subsegment coloring (ascending aorta, red; arch, pink; descending aorta, cyan). Brachiocephalic trunk (red) and left subclavian artery (purple) segmentations are visible, serving as anatomical landmarks for delineation of the ascending, arch, and descending subsegments. The full phenotype report organized by global, ascending, arch, and descending measurements is presented in the right panel.

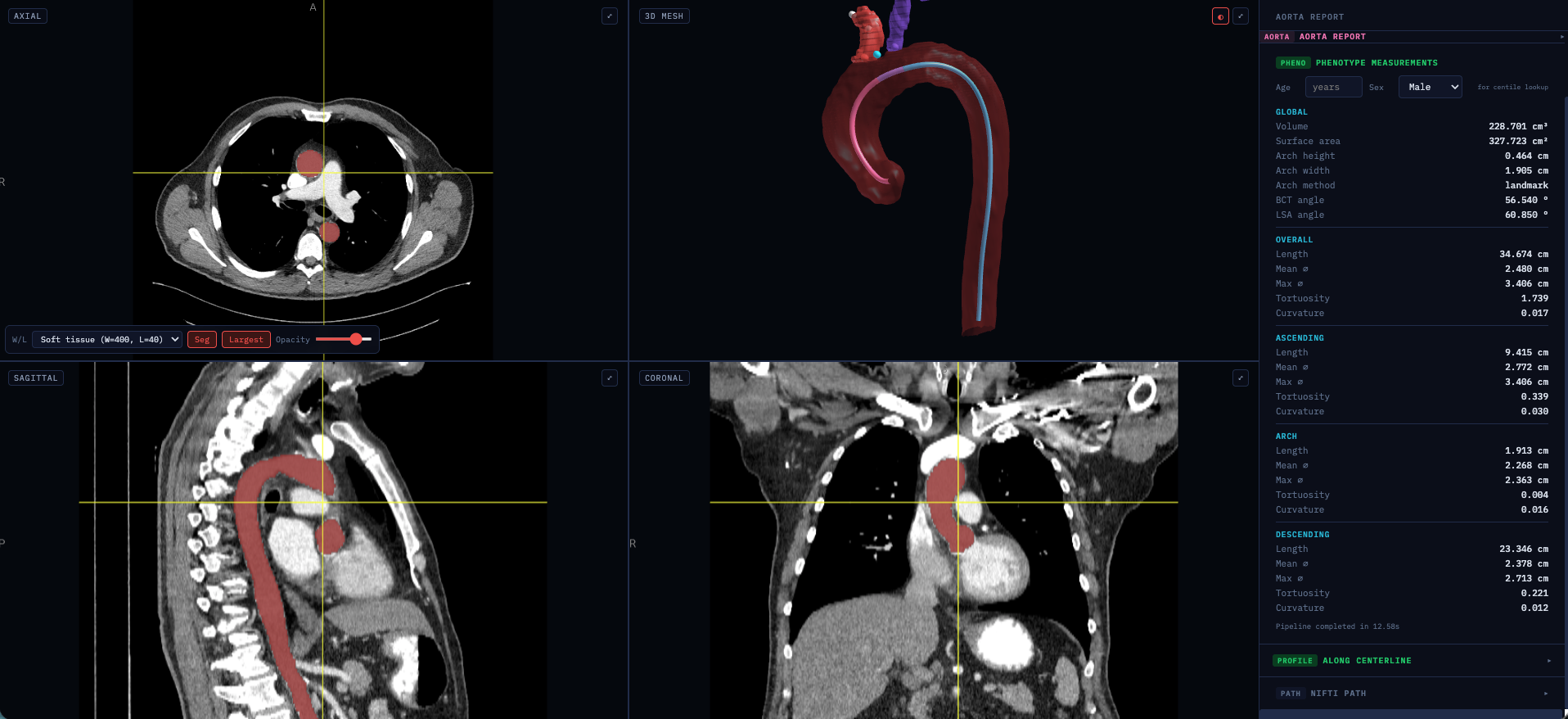

**Figure S2. AGT applied to a non-contrast CT volume.** Screenshot of the AGT visualizer demonstrating geometric phenotyping on a non-contrast CT acquisition. The AGT pipeline operates on the binary aortic segmentation mask rather than image intensities, enabling consistent phenotype extraction independent of contrast administration across the full spectrum of clinical CT protocols.

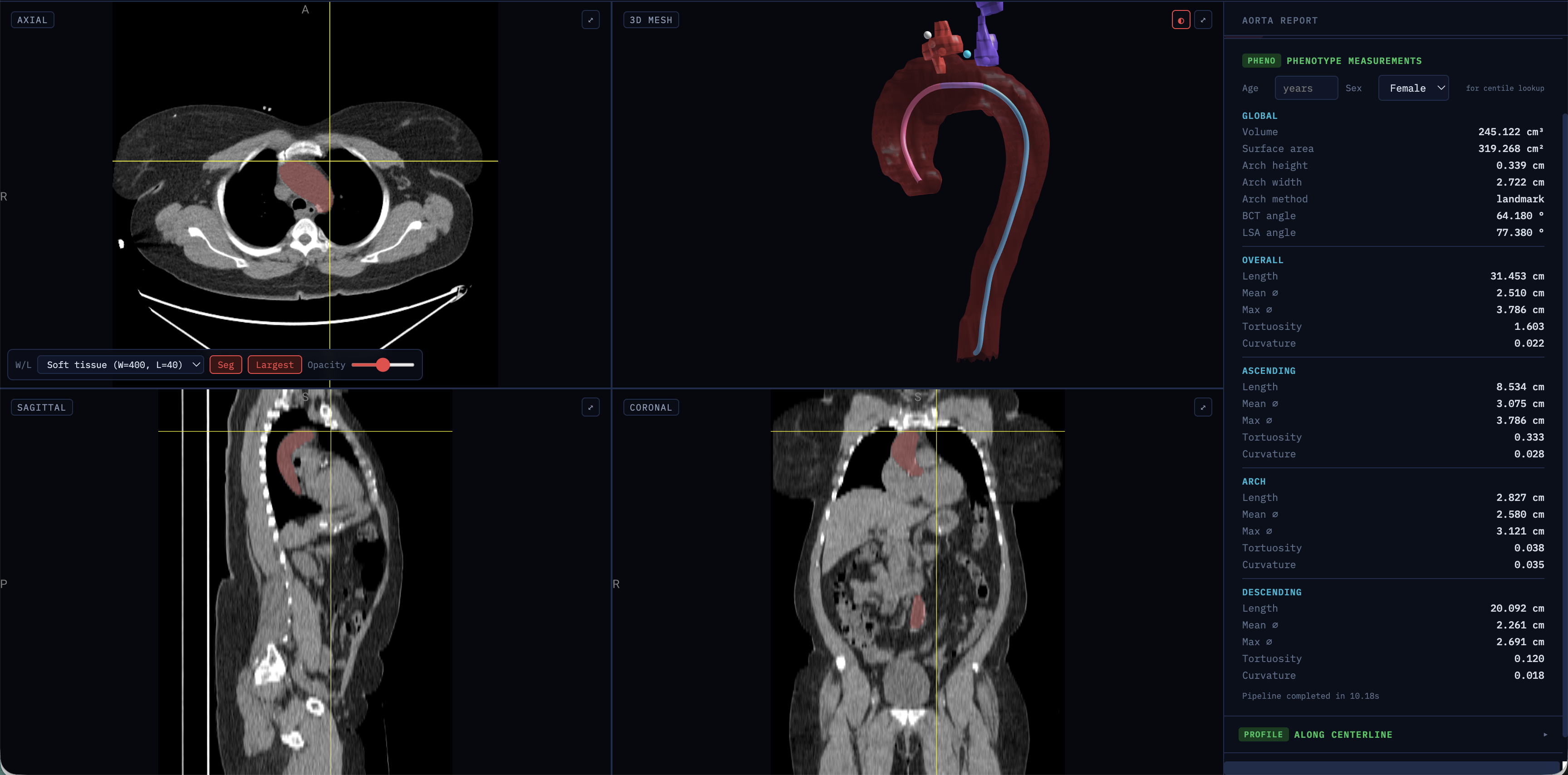

**Figure S3. AGT applied to a participant with aortic atherosclerosis.** Screenshot of the AGT visualizer demonstrating geometric phenotyping in a participant with extensive aortic atherosclerotic calcifications visible on axial (top-left), sagittal (bottom-left), and coronal (bottom-right) views. The field of view extends beyond the thorax into the abdomen, illustrating that the AGT pipeline correctly isolates the thoracic aortic segment regardless of the extent of the acquired volume.

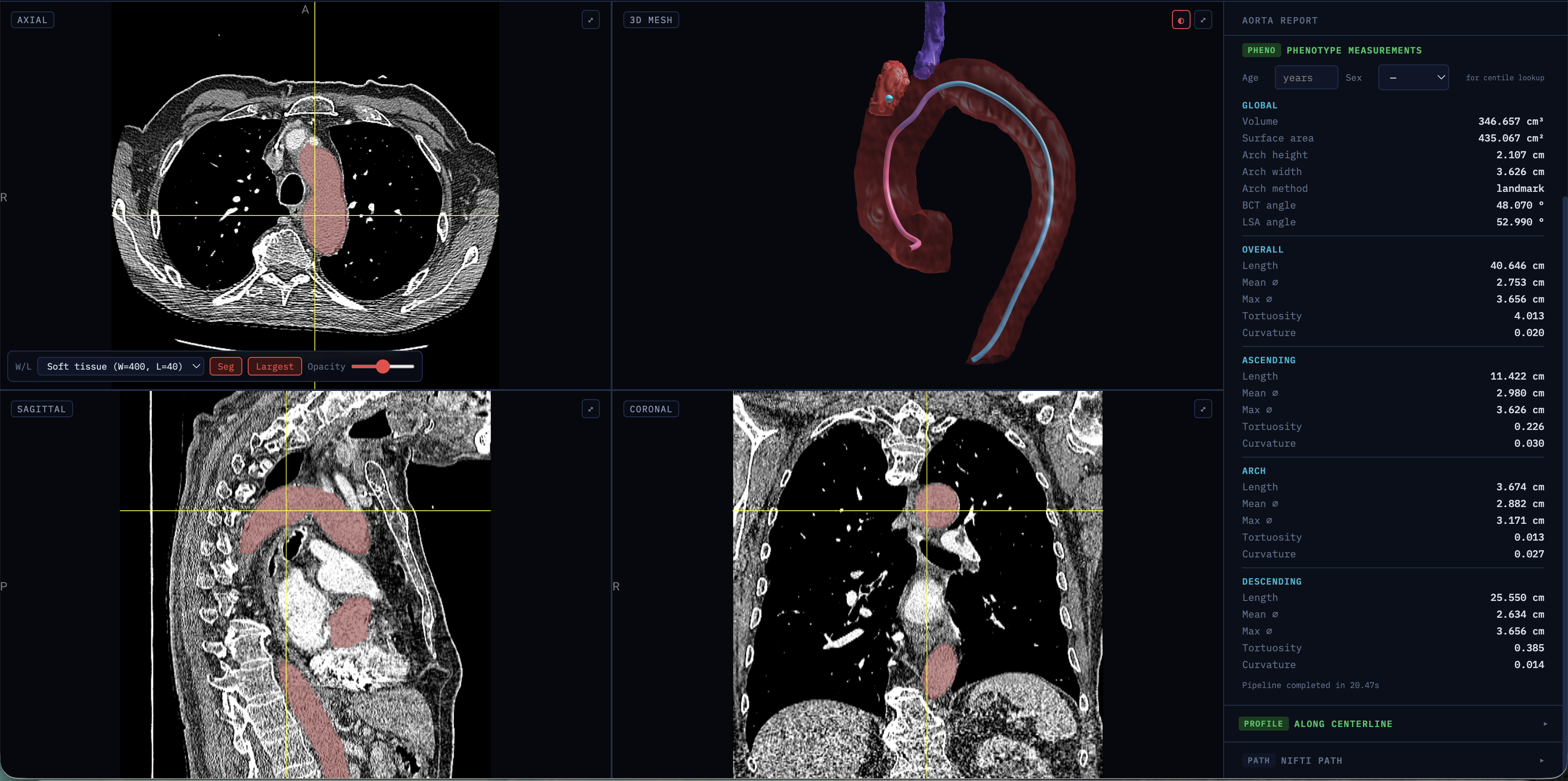

**Figure S4. AGT applied to a participant with aortic atherosclerosis.** Screenshot of the AGT visualizer demonstrating geometric phenotyping in a participant with aortic atherosclerotic calcifications and a markedly tortuous descending aorta, visible in the sagittal (bottom-left) and three-dimensional mesh (top-right) views.

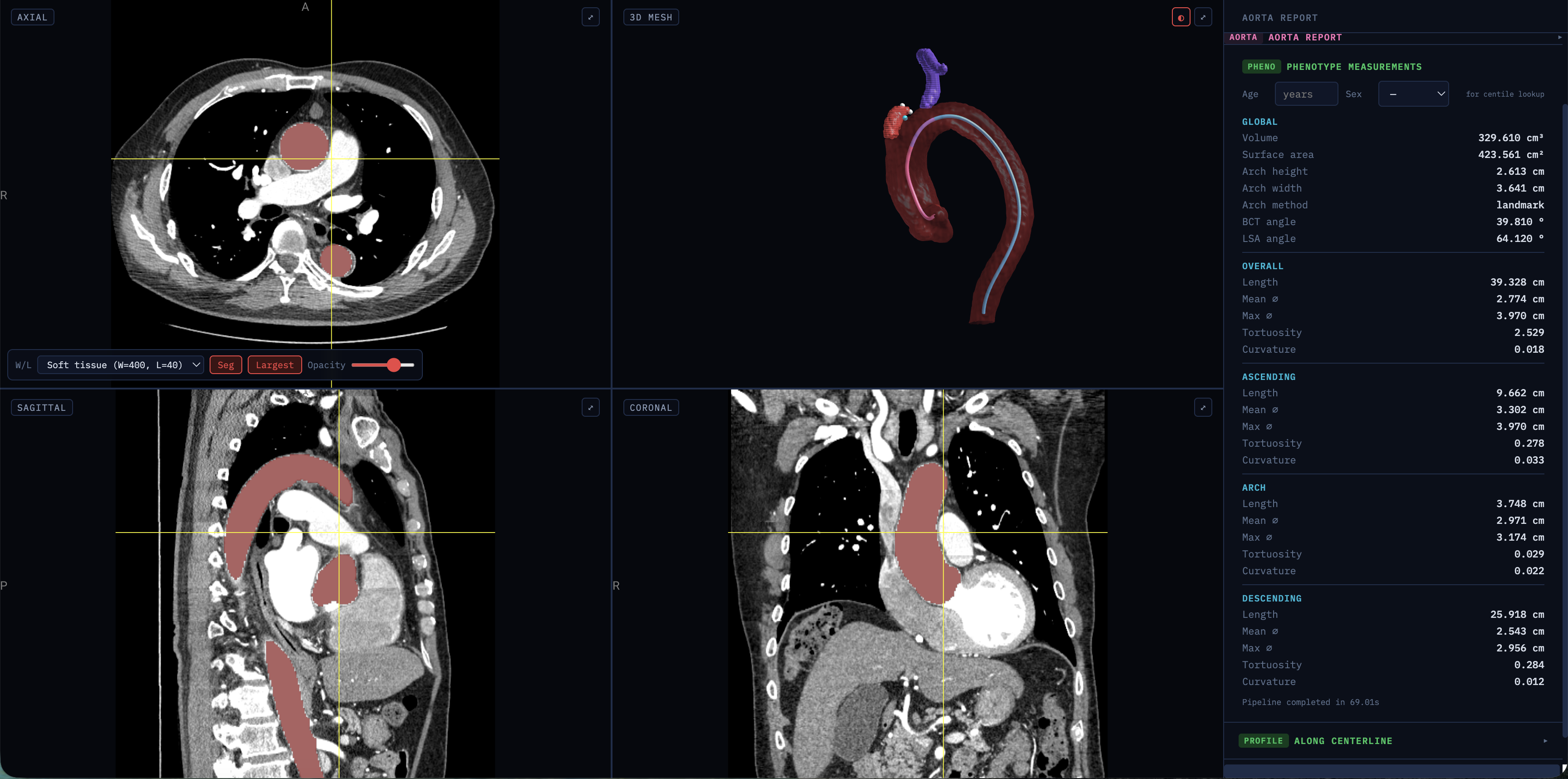

**Figure S5. AGT applied to a participant with ascending aortic aneurysm.** Screenshot of the AGT visualizer demonstrating geometric phenotyping in a participant with an ascending aortic aneurysm, with focal dilation clearly visible on axial (top-left), sagittal (bottom-left), and coronal (bottom-right) views. The segmentation captures the aneurysmal segment, and the three-dimensional mesh (top-right) illustrates the asymmetric bulging of the ascending aorta relative to the arch and descending segments.

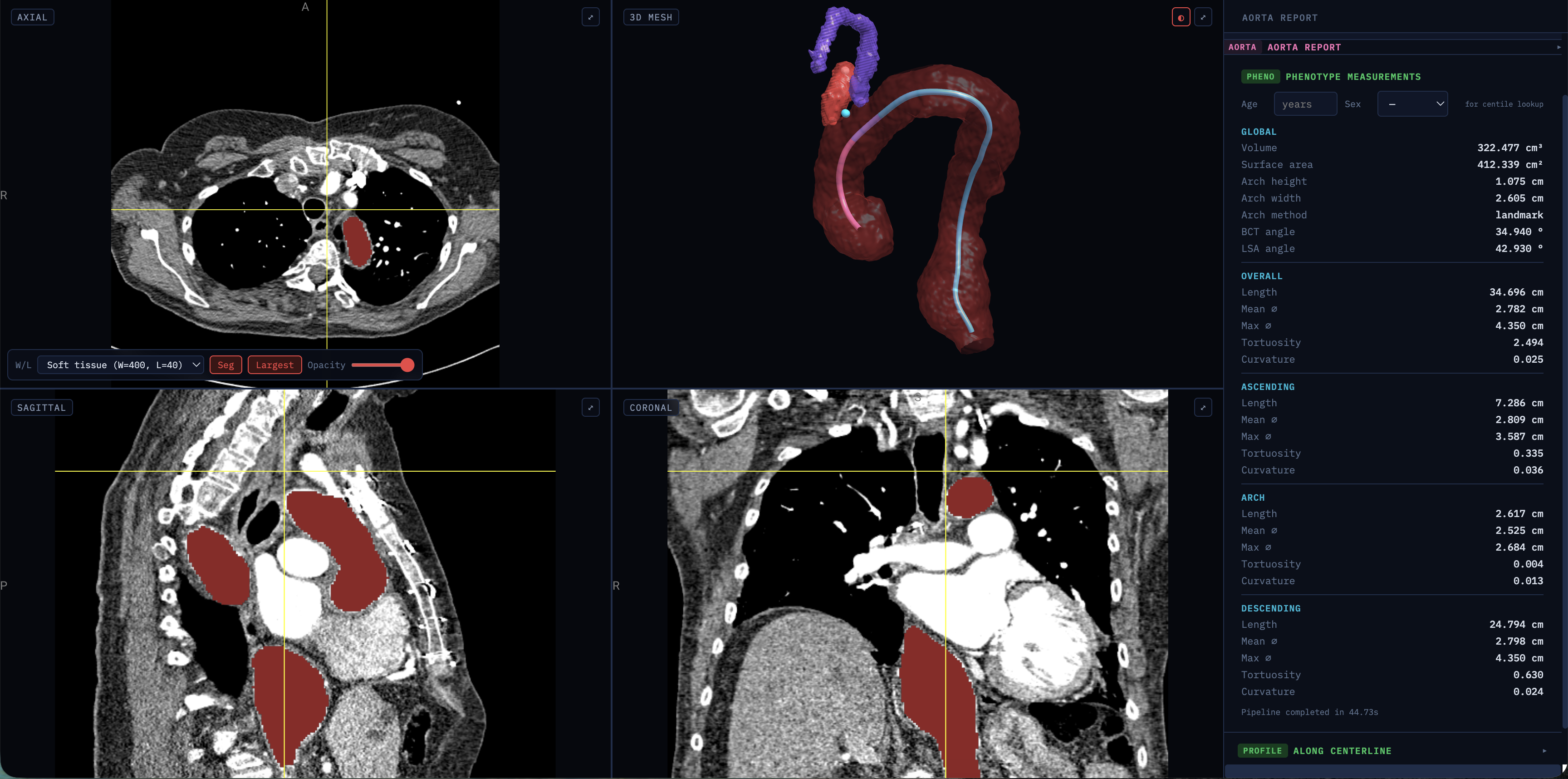

**Figure S6. AGT applied to a participant with descending aortic aneurysm.** Screenshot of the AGT visualizer demonstrating geometric phenotyping in a participant with a descending aortic aneurysm, with focal dilation visible on the sagittal (bottom-left) and coronal (bottom-right) views. The three-dimensional mesh (top-right) illustrates the focal nature of the descending aneurysmal dilation.

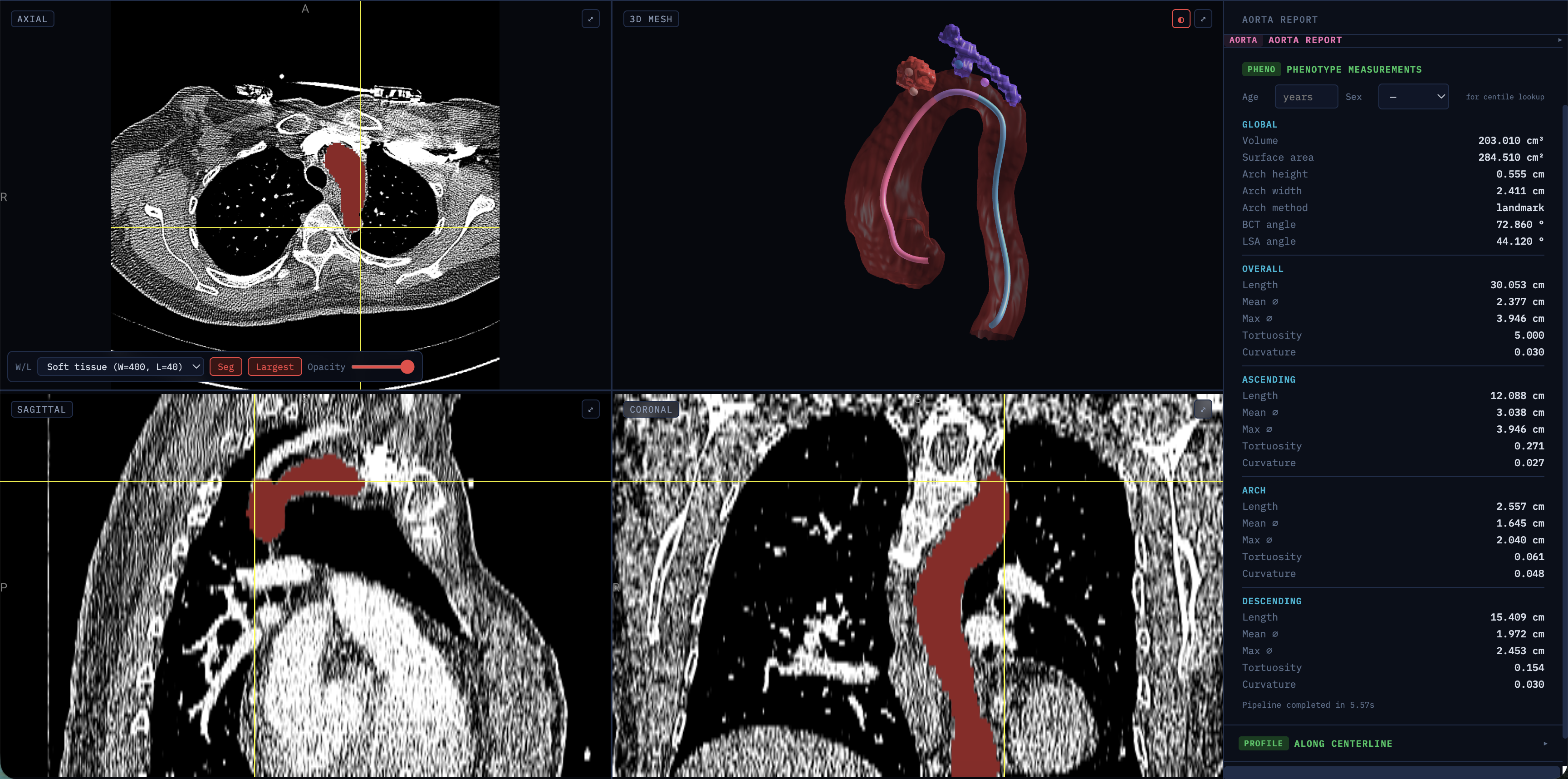

**Figure S7. AGT applied to a participant with aortic coarctation.** Screenshot of the AGT visualizer demonstrating geometric phenotyping in a participant with aortic coarctation. The focal narrowing at the aortic isthmus is visible on the sagittal (bottom-left) and coronal (bottom-right) views, with the three-dimensional mesh (top-right) illustrating the characteristic abrupt caliber transition between the arch and descending segments.

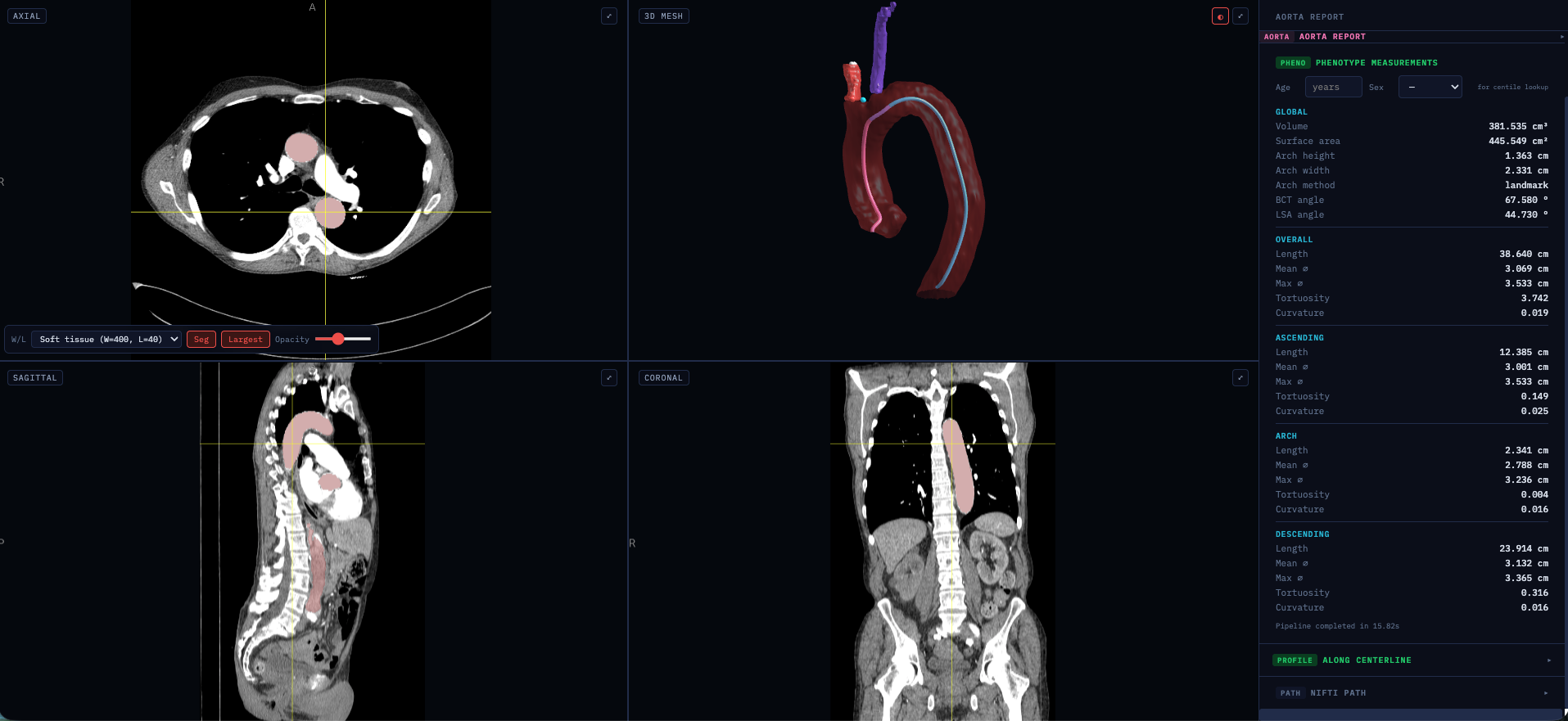

**Figure S8. AGT applied to a participant with aortic dissection.** Screenshot of the AGT visualizer demonstrating geometric phenotyping in a participant with aortic dissection. The aortic segmentation encompasses the full aortic cross-section without distinction between true and false lumens, and the resulting phenotype report reflects the composite geometric distortion characteristic of dissection, including increased total aortic volume (381.5 cm³), and uniformly enlarged diameters across all subsegments.

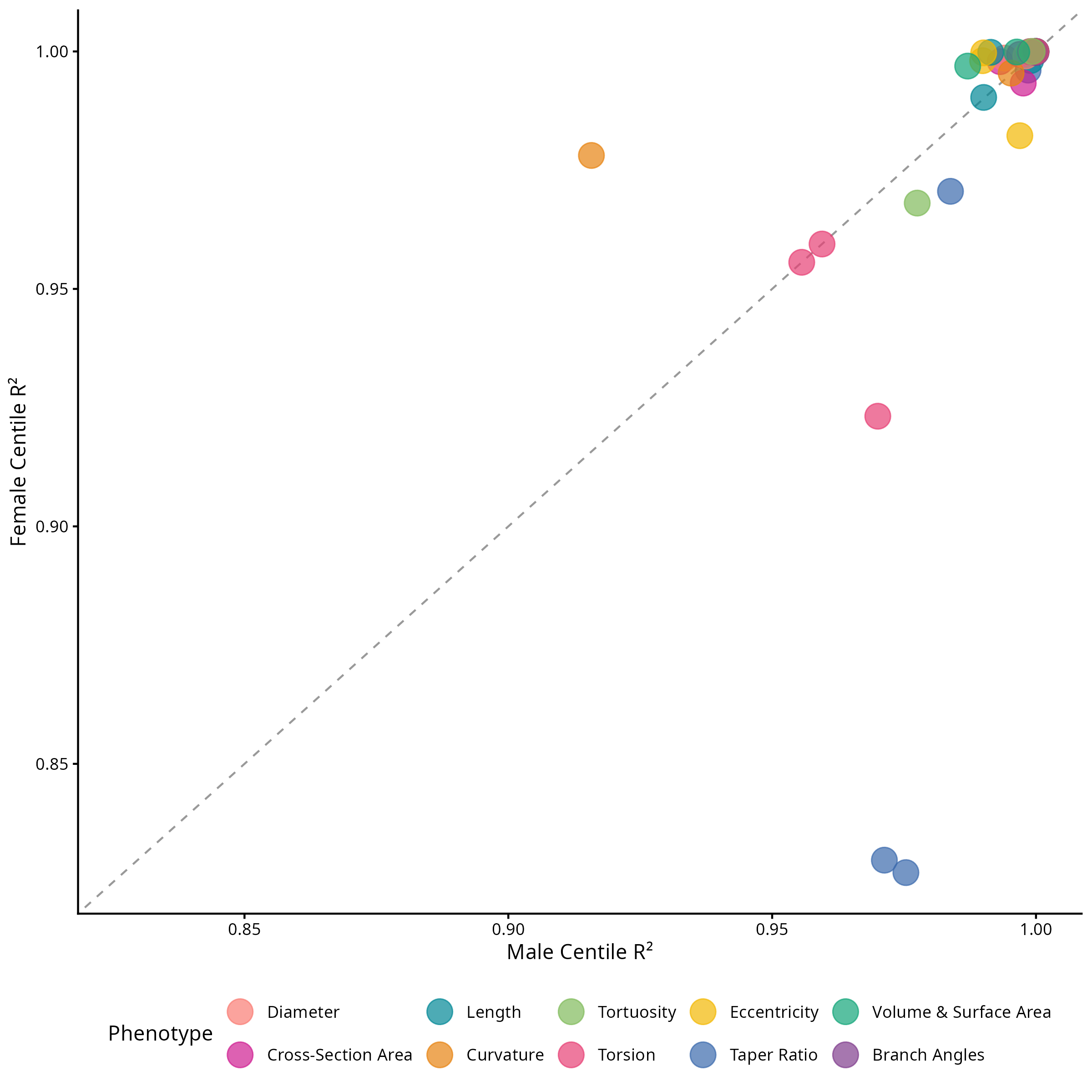

**Figure S9.** **Discovery-replication agreement of median centile curves.** Sex-specific coefficient of determination (R^2^) between the 50-50 discovery and replication splits, with the male R^2^ (x-axis) plotted against the female R^2^ (y-axis) for each AGP, colored by phenotype category. The dashed line indicates the line of identity (y = x), along which the male and female discovery-replication R^2^ values are equal. Centile curves agreed closely across the splits, with R^2^ > 0.95 in 37 of 38 phenotypes for males and 35 of 38 for females (mean R^2^=0.990 male, 0.983 female). The lowest agreement was observed among higher-order phenotypes (taper ratio, curvature, and torsion).

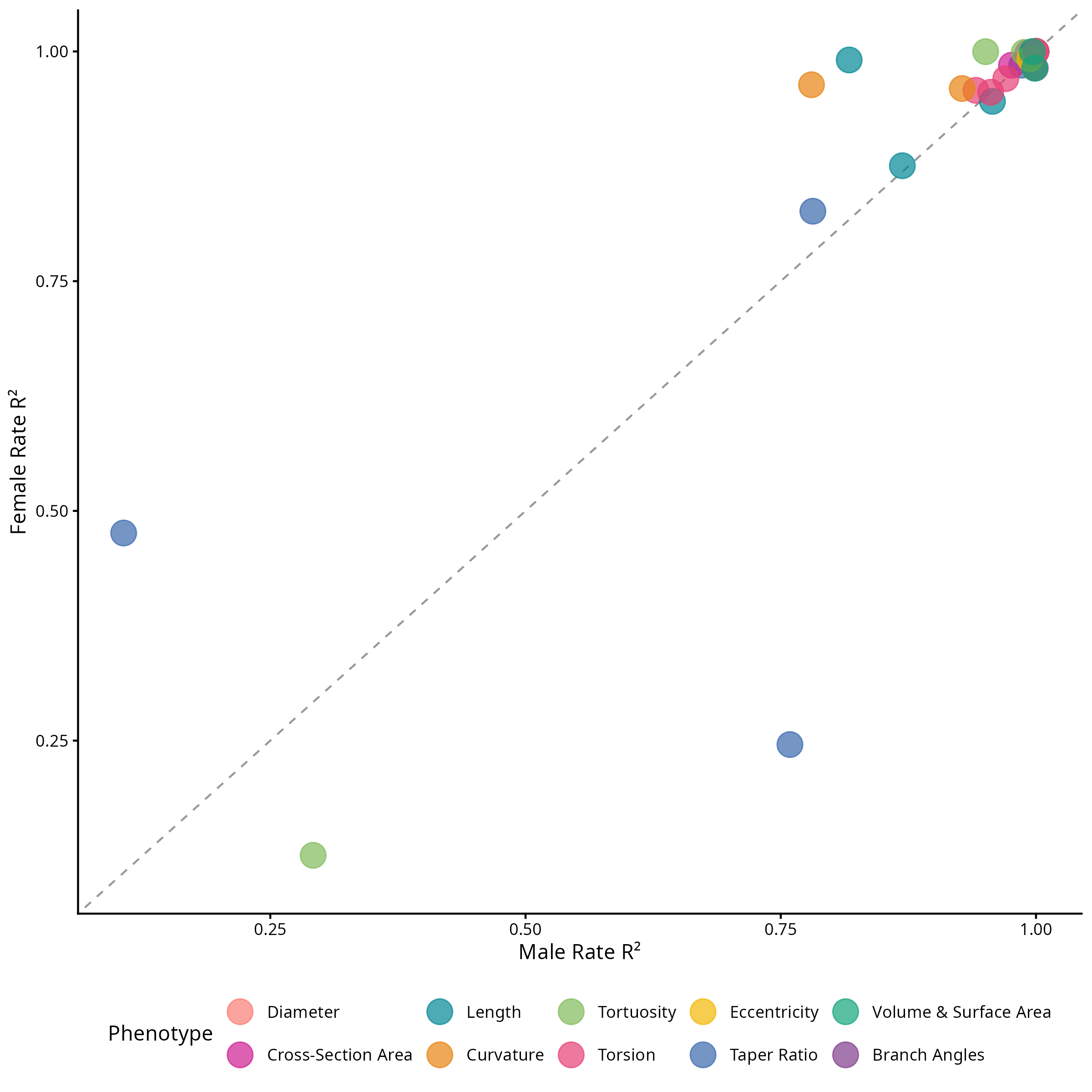

**Figure S10.** Discovery-replication agreement of rate-of-change curves. Sex-specific coefficient of determination (R^2^) between the 50-50 discovery and replication splits for the rate-of-change curves (first derivative of the median centile), with the male R^2^ (x-axis) plotted against the female R^2^ (y-axis) for each AGP, colored by phenotype category. The dashed line indicates the line of identity (y = x), along which the male and female discovery-replication R^2^ values are equal. Rate-of-change curves agreed across the splits, with R^2^ > 0.95 in 29 of 38 phenotypes for males and 32 of 38 for females (mean R^2^=0.922 male, 0.926 female). The lowest agreement was observed among higher-order phenotypes (taper ratio and tortuosity).

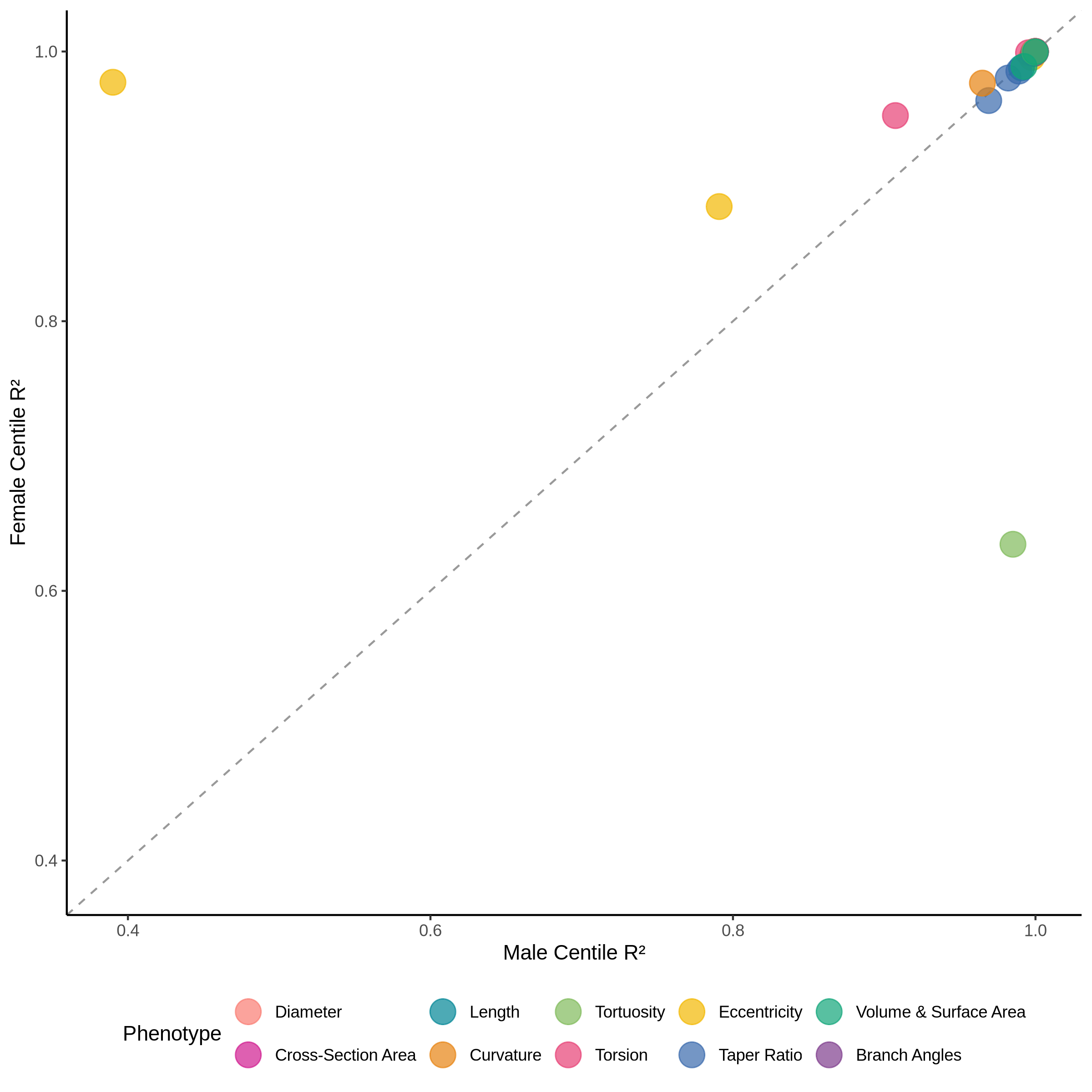

**Figure S11.** **Sensitivity analysis comparing random effect specifications for aging charts**. Sex-specific coefficient of determination (R^2^) for centile curves between the cohort and scanner x cohort random effect GAMLSS model specifications, with the male R^2^ (x-axis) plotted against the female R^2^ (y-axis) for each AGP, colored by phenotype category. The dashed line indicates the line of identity (y = x), along which the male and female R^2^ values are equal. Centile curves agreed closely between specifications, with R^2^ > 0.95 in 35 of 38 phenotypes for males and 36 of 38 for females (mean R^2^=0.972 male, 0.982 female). The lowest agreement was observed among higher-order phenotypes (eccentricity and tortuosity).

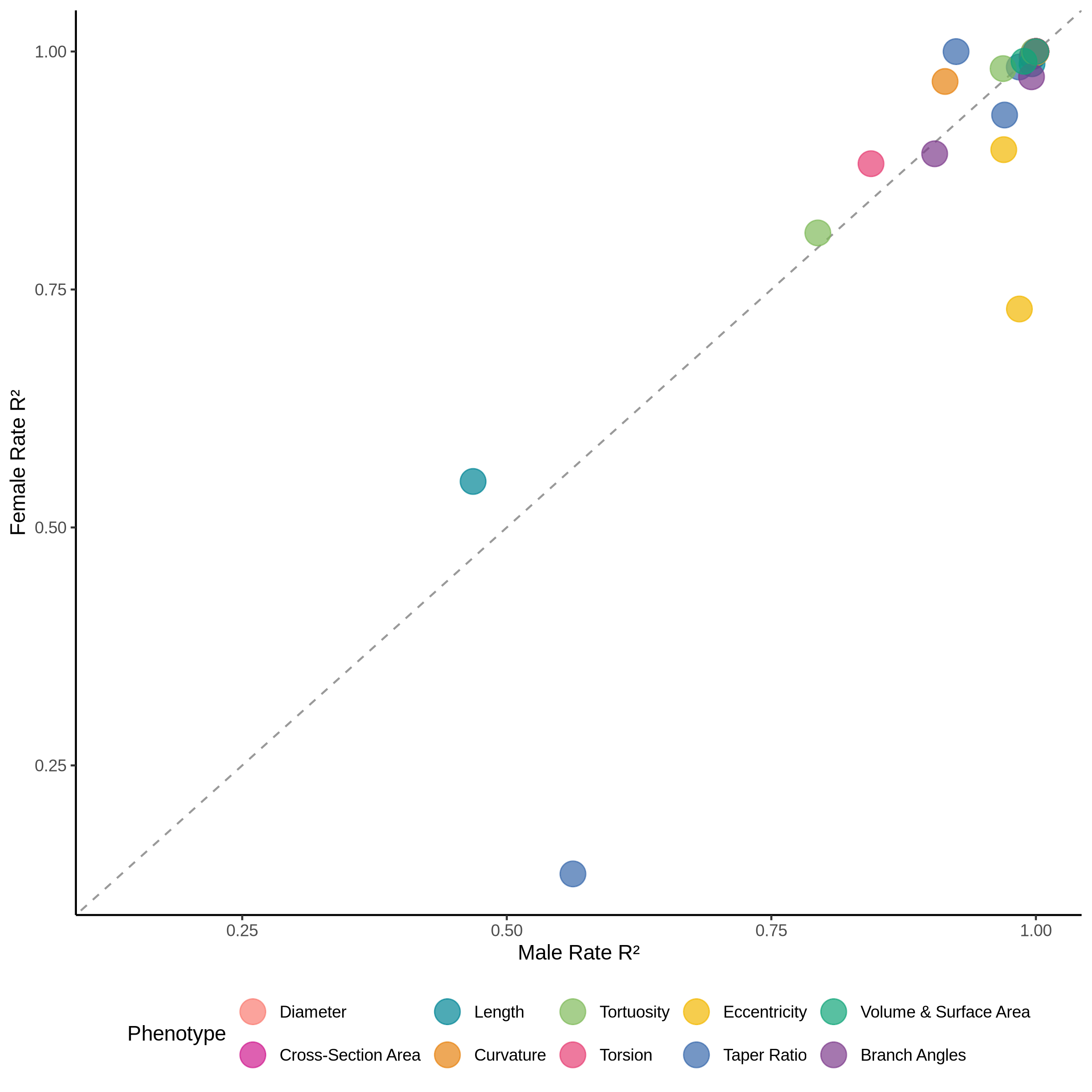

**Figure S12. Sensitivity analysis comparing random effect specifications for rate-of-change curves**. Sex-specific coefficient of determination (R^2^) for rate-of-change curves (first derivative of the median centile) between the cohort and scanner x cohort random effect GAMLSS model specifications, with the male R^2^ (x-axis) plotted against the female R^2^ (y-axis) for each AGP, colored by phenotype category. The dashed line indicates the line of identity (y = x), along which the male and female R^2^ values are equal. Rate-of-change curves agreed between specifications, with R^2^ > 0.95 in 31 of 38 phenotypes for males and 30 of 38 for females (mean R²=0.954 male, 0.939 female). The lowest agreement was observed among higher-order phenotypes (length, taper ratio, and eccentricity).

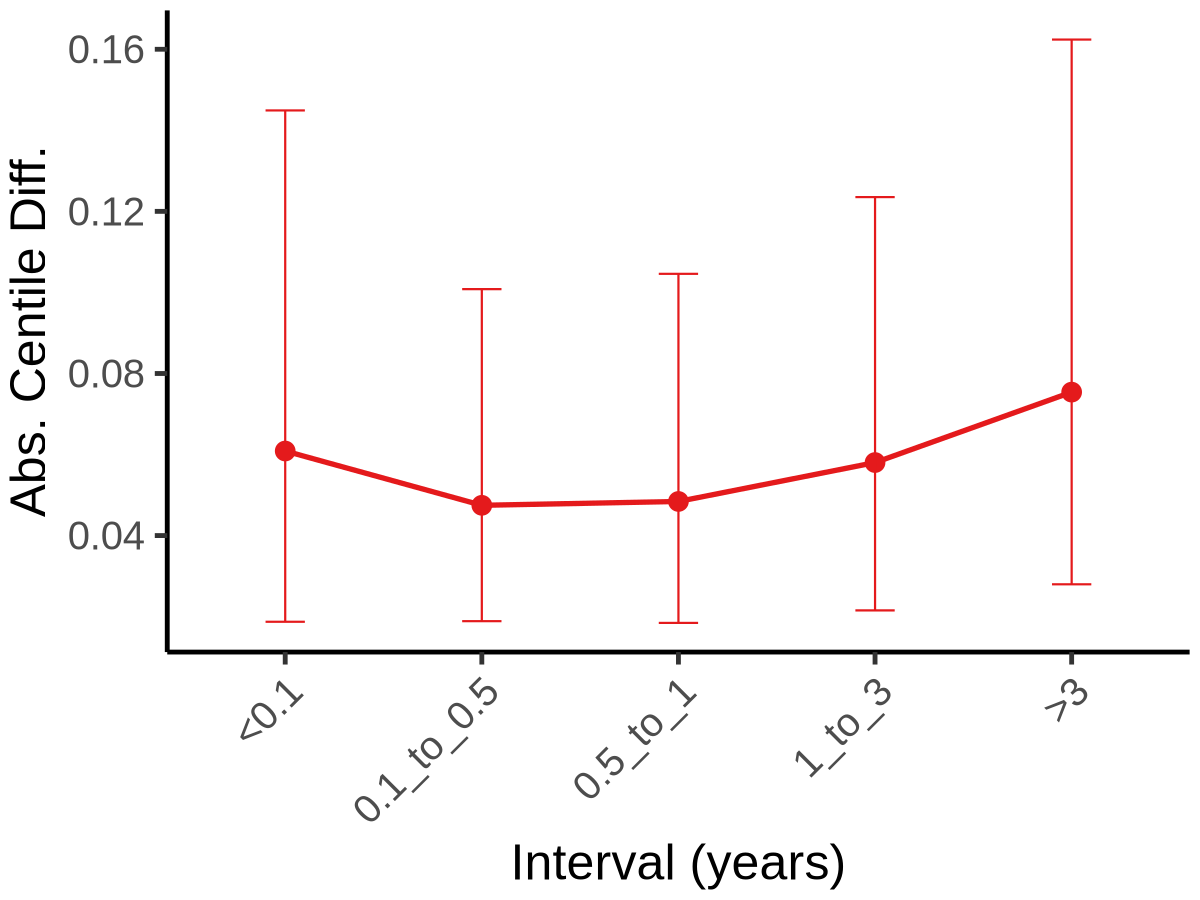

**Figure S13.** Longitudinal stability of aortic volume centile assignment. Median absolute centile difference between paired consecutive CT studies from participants with two or more studies in PMBB or CT-RATE, stratified by interval between studies (years). Points represent medians and error bars indicate the interquartile range. Differences are stable across intervals up to one year, reflecting the cardiac-cycle-dependent measurement floor on non-gated CT, with progressive degradation at longer intervals (1-3 years and >3 years) consistent with cumulative biological drift in aortic volume being captured by the centile framework on top of the cardiac-cycle noise floor.

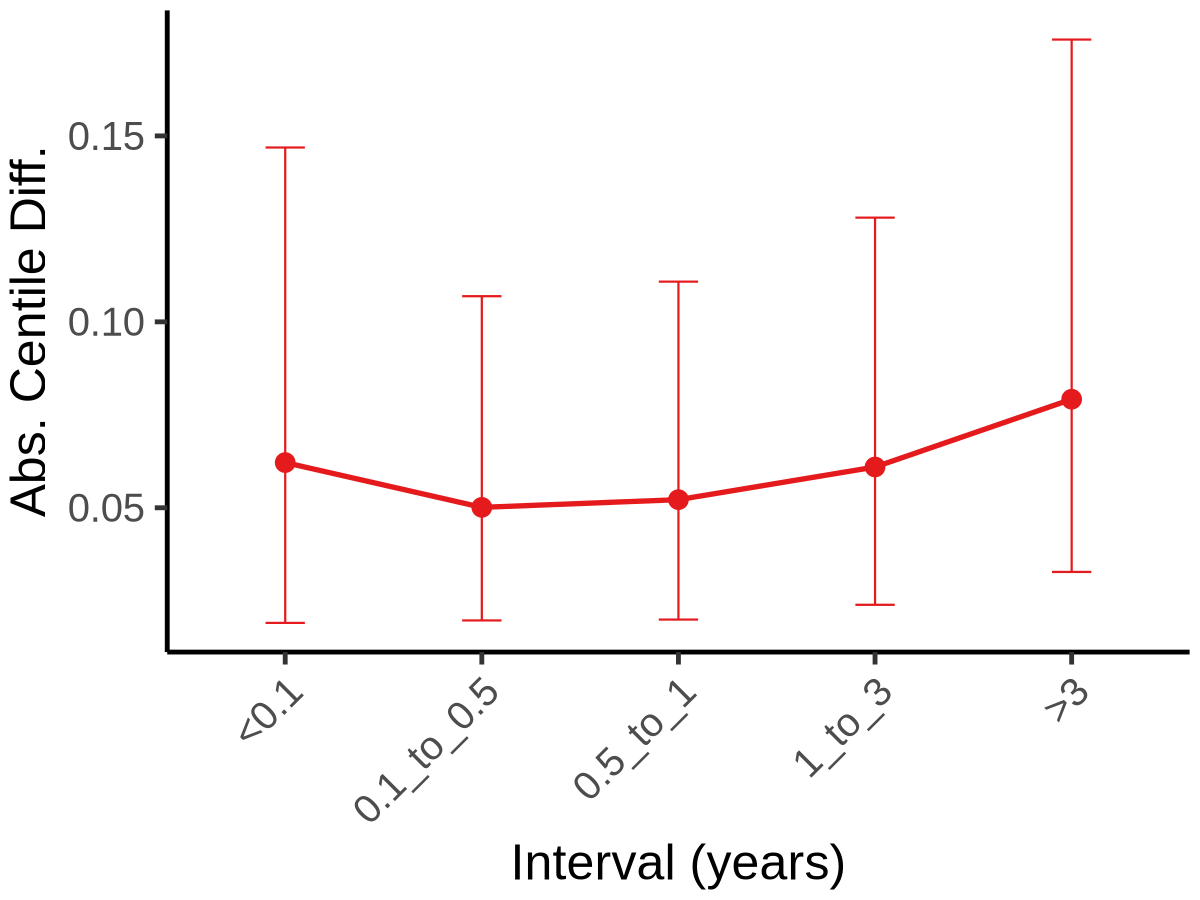

**Figure S14.** Longitudinal stability of aortic surface area centile assignment. Median absolute centile difference between paired consecutive CT studies from participants with two or more studies in PMBB or CT-RATE, stratified by interval between studies (years). Points represent medians and error bars indicate the interquartile range (IQR).

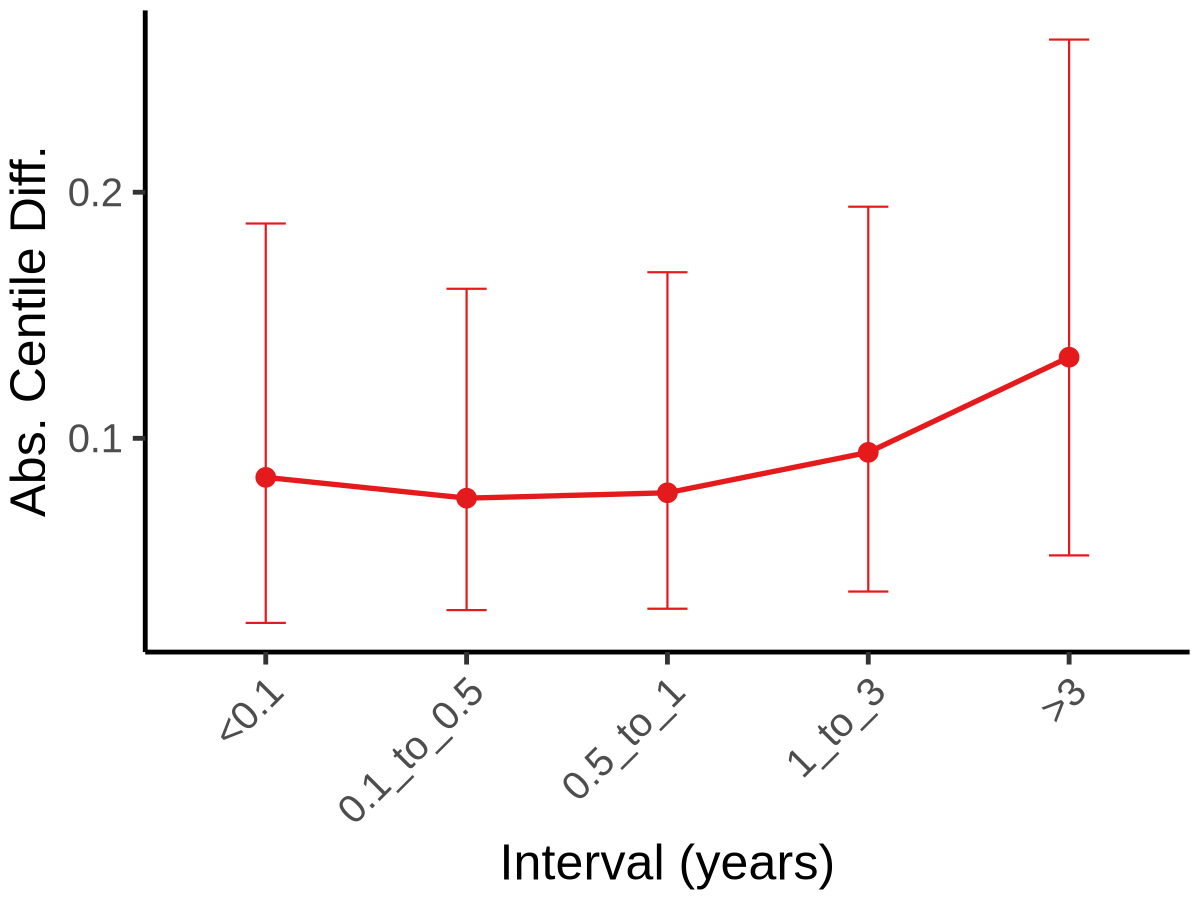

**Figure S15.** Longitudinal stability of aortic cross-sectional area centile assignment. Median absolute centile difference between paired consecutive CT studies from participants with two or more studies in PMBB or CT-RATE, stratified by interval between studies (years). Points represent medians and error bars indicate the interquartile range (IQR).

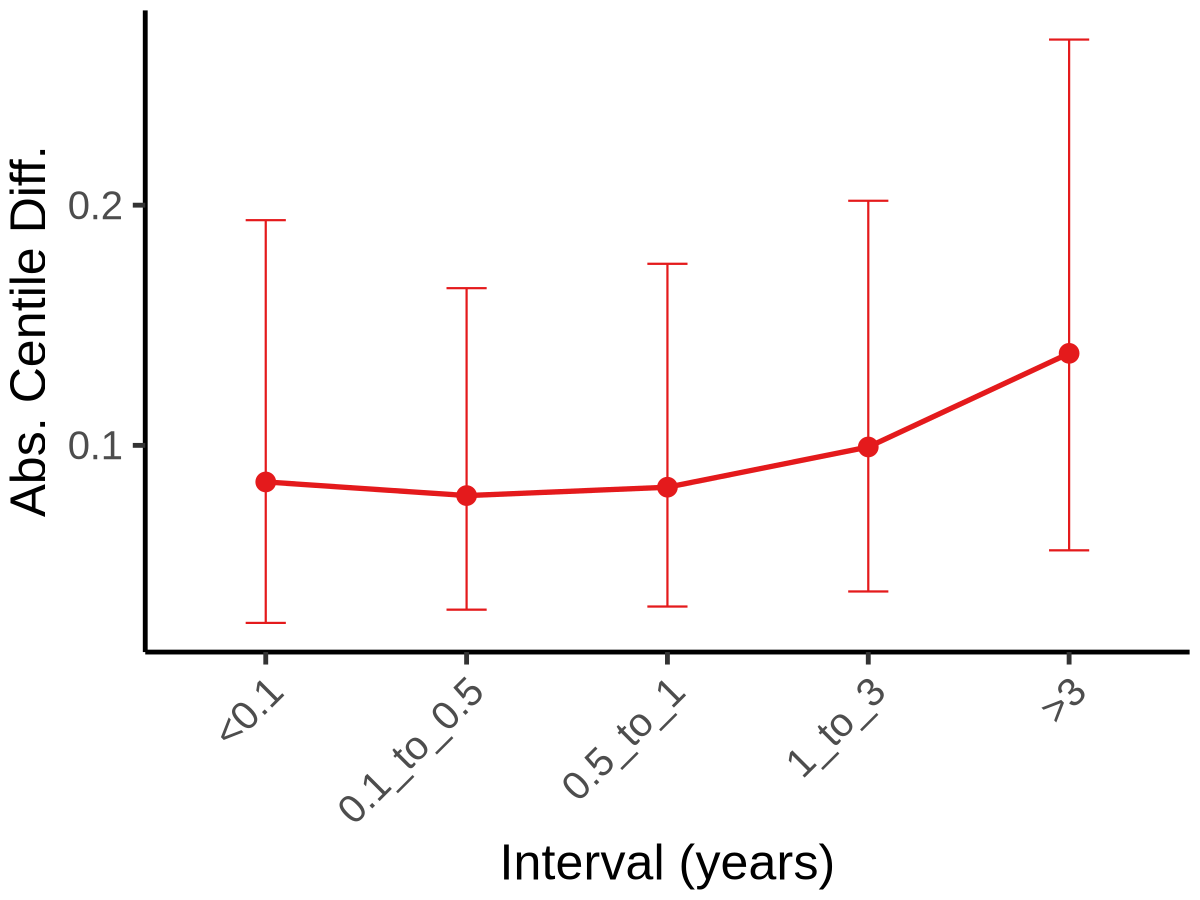

**Figure S16.** Longitudinal stability of thoracic aortic diameter. Median absolute centile difference between paired consecutive CT studies from participants with two or more studies in PMBB or CT-RATE, stratified by interval between studies (years). Points represent medians and error bars indicate the interquartile range (IQR).

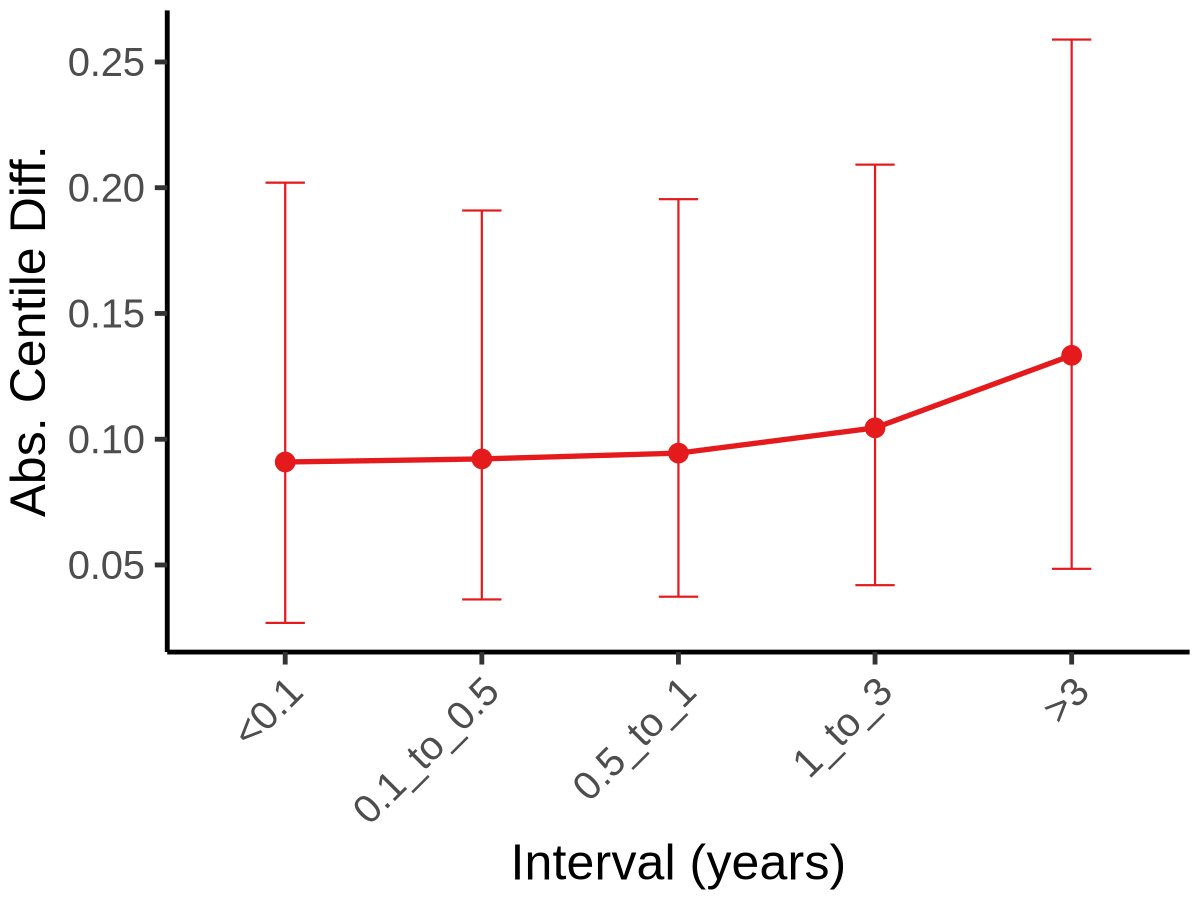

**Figure S17.** Longitudinal stability of thoracic aortic centerline length. Median absolute centile difference between paired consecutive CT studies from participants with two or more studies in PMBB or CT-RATE, stratified by interval between studies (years). Points represent medians and error bars indicate the interquartile range (IQR).

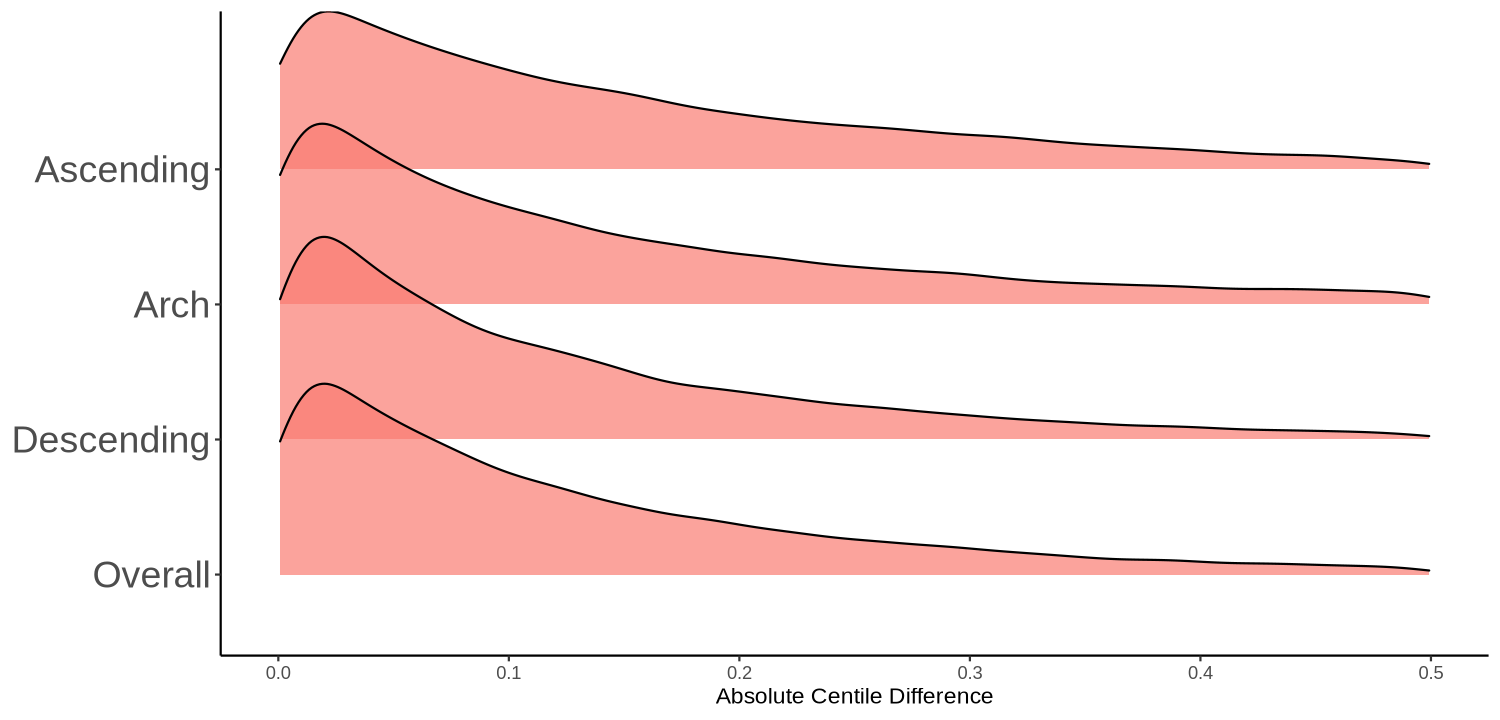

**Figure S18.** Longitudinal stability of thoracic aortic diameter, stratified by aortic subsegments. Density plot visualization of median absolute centile difference between paired consecutive CT studies from participants with two or more studies in PMBB or CT-RATE, stratified thoracic aortic subsegment.

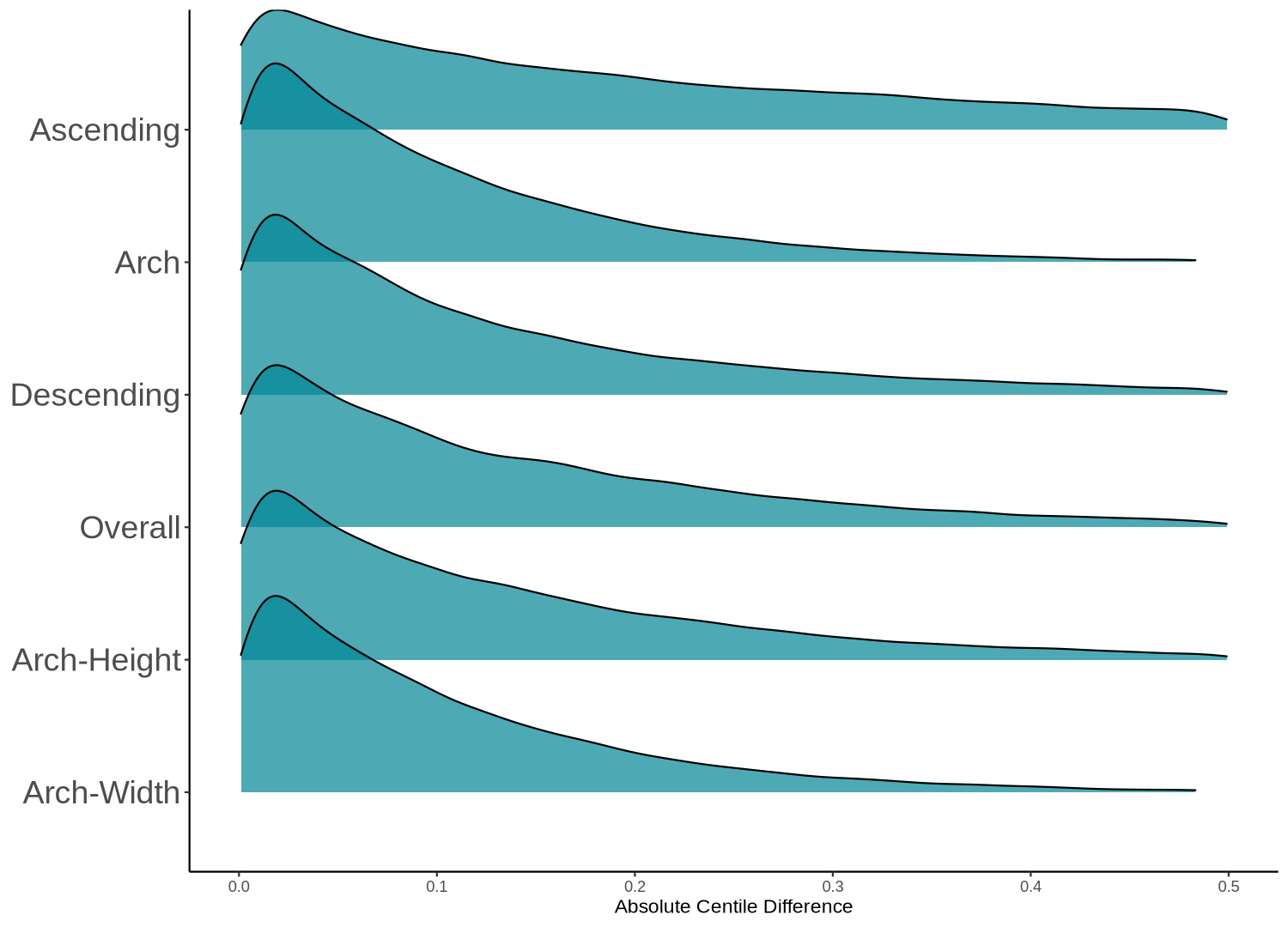

**Figure S19.** Longitudinal stability of thoracic aortic length, stratified by aortic subsegments. Two additional, region-specific measurements for the curvilinear segment of the thoracic aortic arch are included, namely arch-height and arch-width. Density plot visualization of median absolute centile difference between paired consecutive CT studies from participants with two or more studies in PMBB or CT-RATE, stratified by thoracic aortic subsegment.

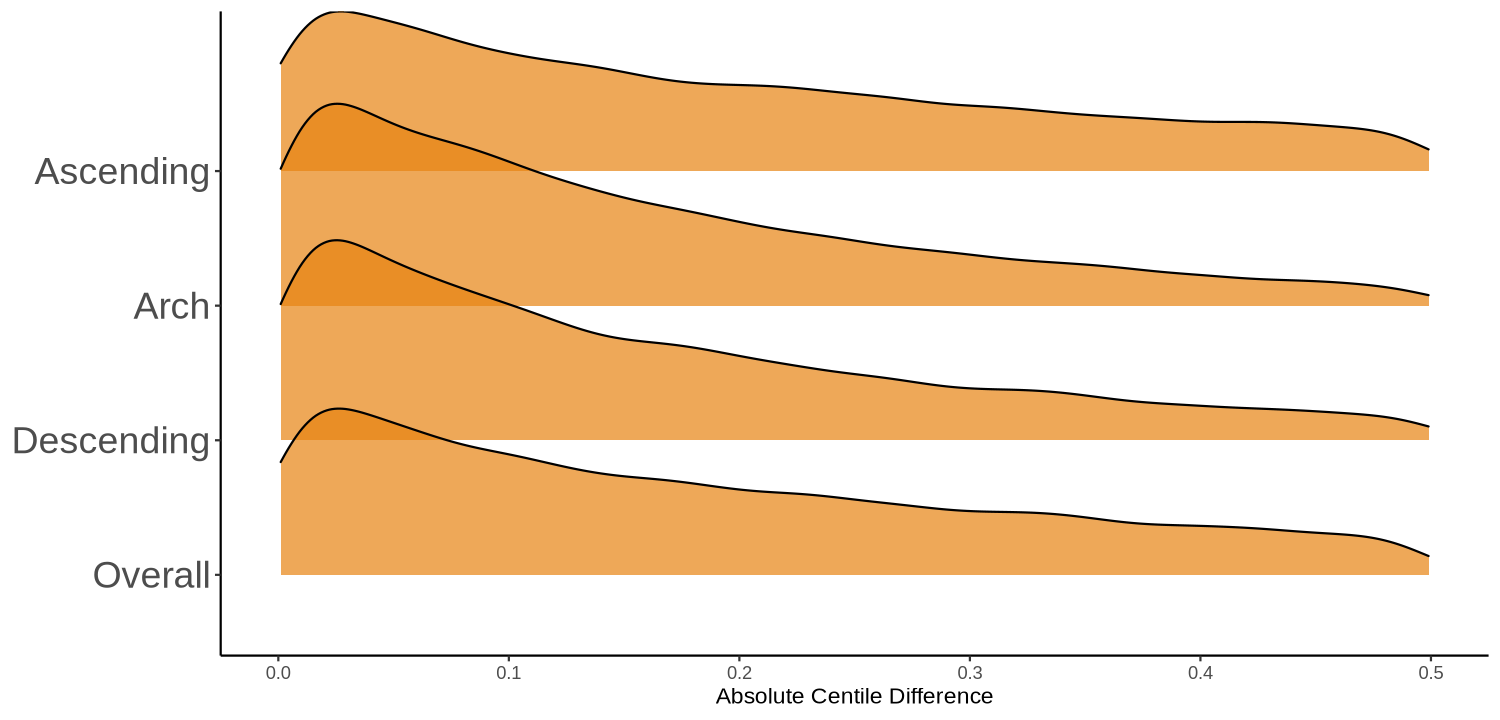

**Figure S20.** Longitudinal stability of aortic curvature, a second-order aortic geometric phenotype, stratified by aortic subsegments. Density plot visualization of median absolute centile difference between paired consecutive CT studies from participants with two or more studies in PMBB or CT-RATE, stratified by thoracic aortic subsegment.

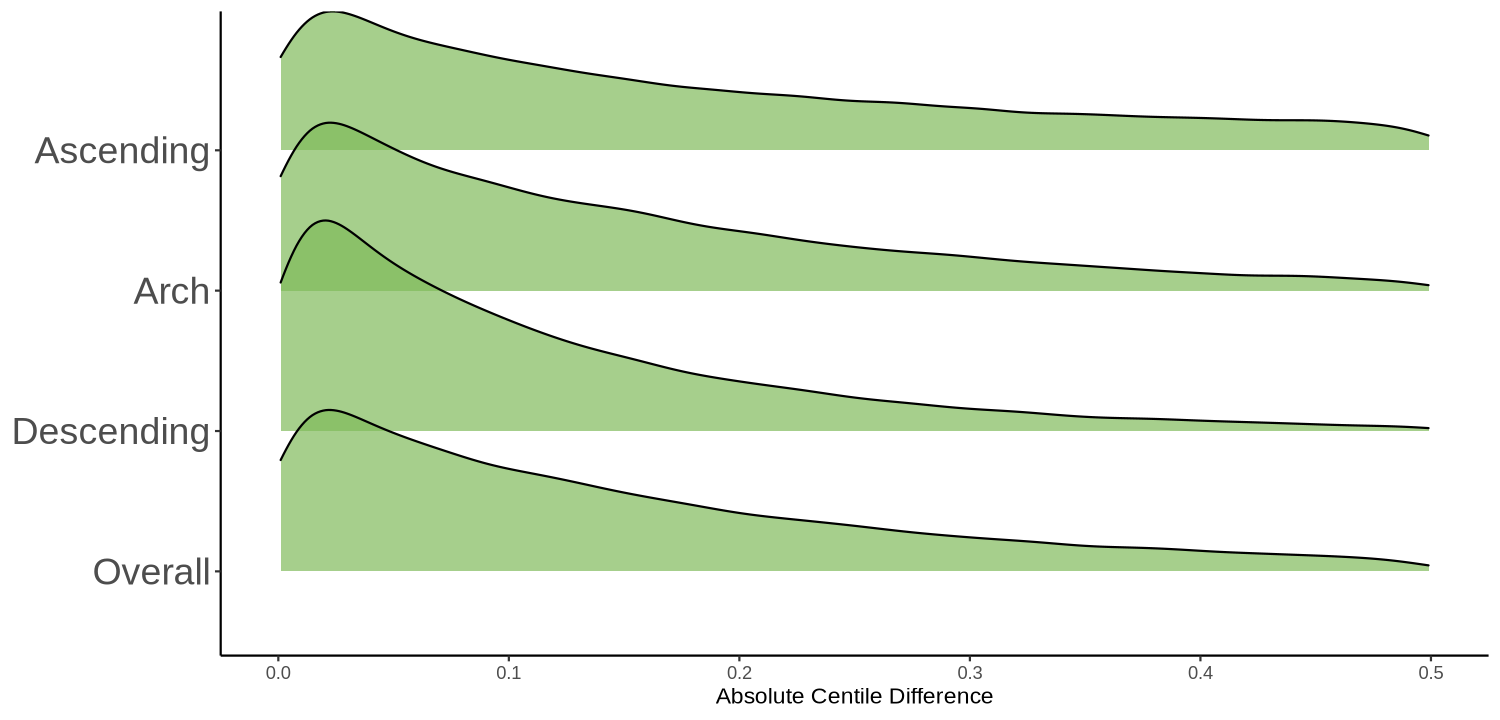

**Figure S21.** Longitudinal stability of aortic tortuosity, a second-order aortic geometric phenotype, stratified by aortic subsegments. Density plot visualization of median absolute centile difference between paired consecutive CT studies from participants with two or more studies in PMBB or CT-RATE, stratified by thoracic aortic subsegment.

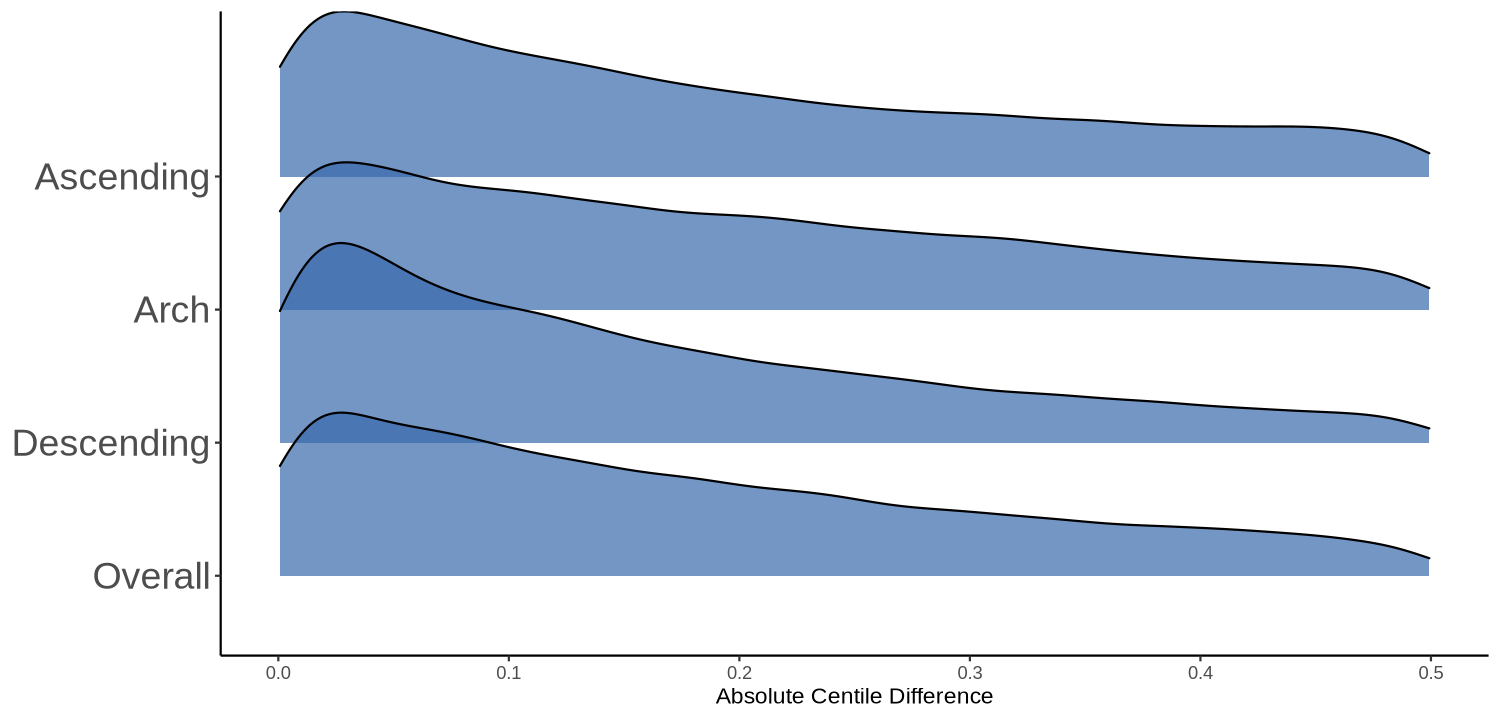

**Figure S22.** Longitudinal stability of aortic taper ratio, a second-order aortic geometric phenotype, stratified by aortic subsegments. Density plot visualization of median absolute centile difference between paired consecutive CT studies from participants with two or more studies in PMBB or CT-RATE, stratified by thoracic aortic subsegment.

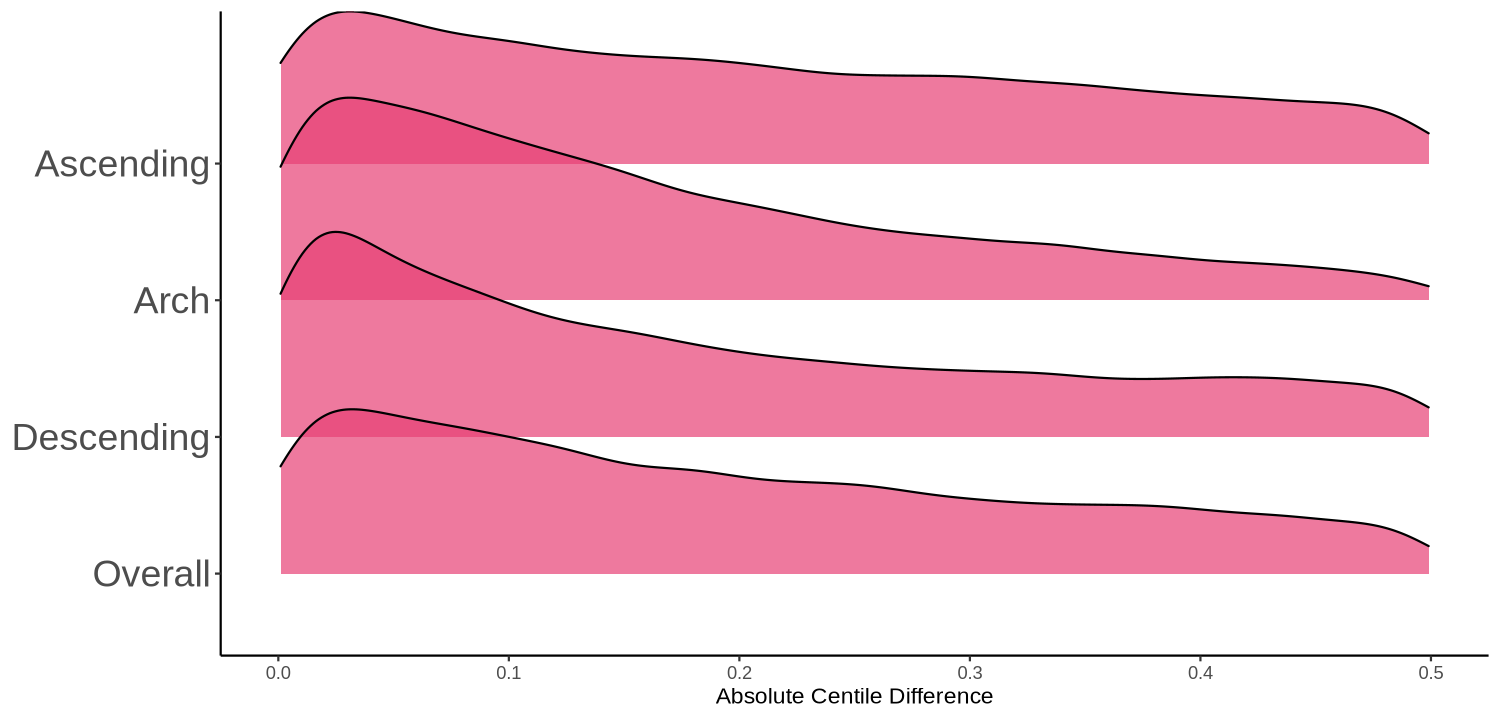

**Figure S23.** Longitudinal stability of aortic torsion, a third-order aortic geometric phenotype, across thoracic aortic subsegments. Density distributions of the median absolute centile difference between paired consecutive CT studies from participants with two or more studies in PMBB or CT-RATE, stratified by thoracic aortic subsegment.

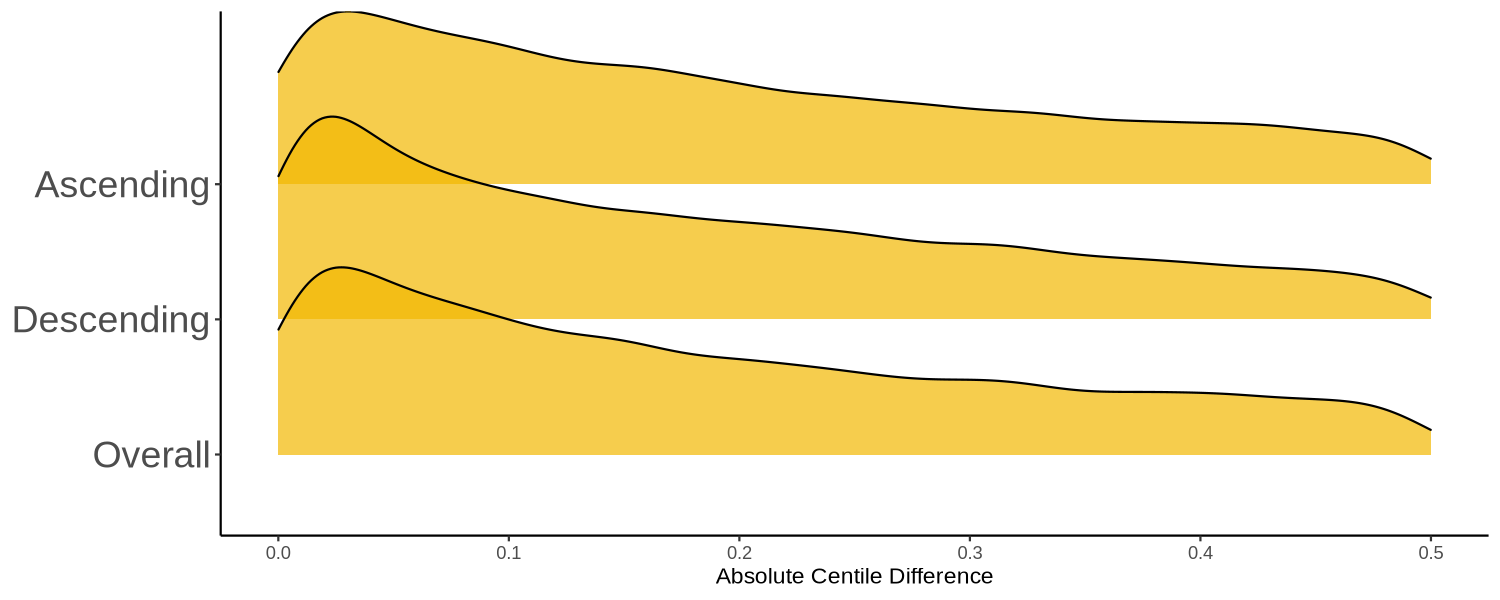

**Figure S24.** Longitudinal stability of aortic eccentricity, a third-order aortic geometric phenotype, across thoracic aortic subsegments. Density distributions of the median absolute centile difference between paired consecutive CT studies from participants with two or more studies in PMBB or CT-RATE, stratified by thoracic aortic subsegment. Eccentricity is reported for the ascending aorta, descending aorta, and the aorta overall, but not for the aortic arch, where the oblique orientation of the vessel relative to the axial imaging plane degrades cross-sectional shape estimation.

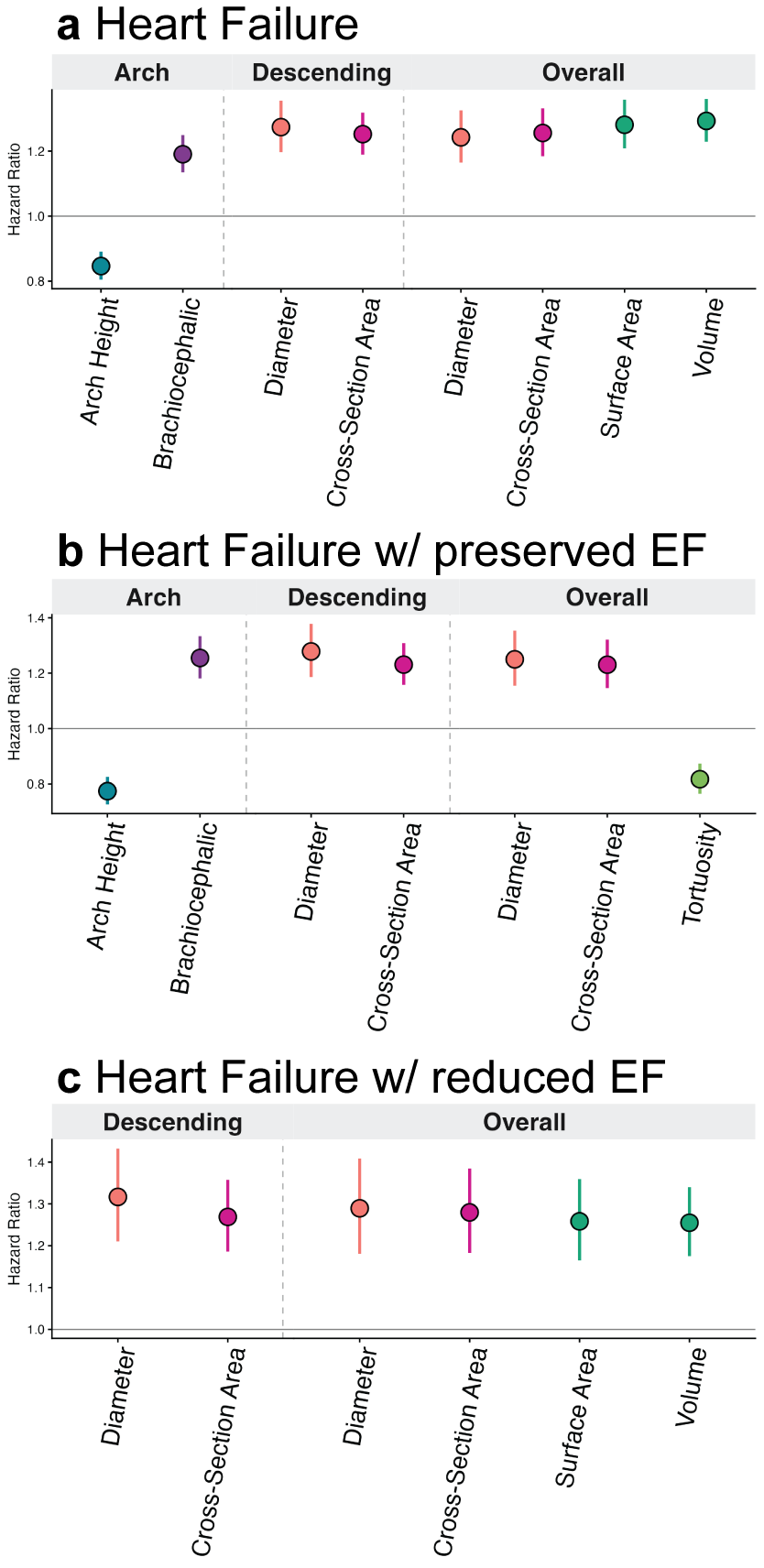

**Figure S25.** Forest plot of significant (*P* < 1.018x10^-6^) aortic geometric phenotypes (AGPs) with hazard ratios and 95% confidence intervals associated with incident heart failure events and their subtypes: (**a**) Heart failure. (**b**) Heart failure with preserved ejection fraction (HFpEF). (**c**) Heart failure with reduced ejection fraction (HFrEF).
